# Sleep-like slow waves during wakefulness uncover a malignant form of Parkinson’s disease

**DOI:** 10.1101/2025.08.29.25334276

**Authors:** Fosco Bernasconi, Thomas Andrillon, Juan Carlos Farah, Isabelle Arnulf, Sophie Schwartz, Jevita Potheegadoo, Saul Martinez-Horta, Javier Pagonabarraga, Jaime Kulisevsky, Olaf Blanke

## Abstract

Slow waves during sleep are fundamental for neural homeostasis, metabolic regulation, and waste clearance, and are known to be altered in neurodegenerative diseases. Sleep-like slow waves (SLSW) have been observed in awake healthy individuals, where they are linked to fluctuating vigilance and mind-wandering. Whether SLSW are altered in Parkinson’s disease (PD), and whether such alterations contribute to psychiatric or cognitive complications, remains unknown. In a cohort of 84 non-demented PD patients and 30 healthy older adults, we found that SLSW occurred more frequently in PD. Hallucinatory traits in PD were associated with higher SLSW amplitude, especially over fronto-central regions, and the extent of SLSW alterations was associated with the severity of hallucinatory burden. Moreover, more prominent SLSW predicted greater cognitive impairment. We conclude that, while slow waves during sleep have positive physiological functions, exacerbated diurnal SLSW may represent reactions related to the PD neuropathology, at the cost of hallucinations and cognitive impairment.

## Introduction

Slow waves during sleep are a characteristic feature of deep sleep, especially during non-rapid eye movement (NREM) sleep, and have been argued to play a crucial role in homeostasis (Huber et al., 2006; Tononi & Cirelli, 2014; Vyazovskiy et al., 2008, 2011; Vyazovskiy & Harris, 2013), including metabolic regulation and clearance of metabolic waste products from the brain (Fultz et al., 2019). At the neural level, slow waves during sleep correspond to slow fluctuations of neuronal membrane potentials, alternating between “on-periods” (spiking neural activity; corresponding to the positive deflection in the slow wave in scalp recordings) and “off-periods” (neural silence; corresponding to the negative deflection in the slow wave) (Fiorillo et al., 2019; Neske, 2016; Steriade, 2003; Vyazovskiy & Harris, 2013). Extended off-periods of neural silence have also been associated with behavioral changes such as sensory disconnection, unresponsiveness, and loss of consciousness (Andrillon et al., 2016; Steriade, 2003; Tononi & Massimini, 2008).

Altered sleep and altered slow waves are consistently observed in neurodegenerative diseases. For instance, decreased slow waves during sleep have been linked to impaired protein clearance (Ju et al., 2014, 2017; Kang et al., 2009; Lee et al., 2020; Sprecher et al., 2017). In a PD animal model, enhanced slow waves improved protein clearance and reduced α-synuclein burden (Morawska et al., 2021), compatible with a potential neuroprotective role of slow waves during sleep (Büchele et al., 2018; Diederich et al., 2009; Minakawa, 2022; Schreiner et al., 2019). Reduced slow waves during sleep in individuals with PD has also been associated with a more malignant form of PD, characterized by greater cognitive impairment, particularly in frontal lobe functions (Schreiner et al., 2021) and a more rapid motor progression (Schreiner et al., 2019). Reduced slow waves during sleep in PD have also been associated with sleep disturbances such as poorer sleep quality, excessive daytime sleepiness (Schreiner et al., 2023), poor sleep efficiency, more frequent awakenings, and decreased total sleep time (Dodet et al., 2024; Iranzo et al., 2024).

Emerging evidence suggests that sleep and wake states exist on a continuum rather than as two strictly separate states (Andrillon & Oudiette, 2023; Nir & Tononi, 2010), and even in fully awake individuals, localized sleep intrusions may occur during wakefulness (Nir et al., 2011; Pigarev et al., 1997; Rector et al., 2009; Sheybani et al., 2023; Vyazovskiy et al., 2011). Moreover, such localized sleep intrusions during wakefulness have electrophysiological signatures resembling slow waves during NREM sleep. These activity patterns have been termed sleep-like slow waves (SLSW) and have been observed in the delta (0.5–4 Hz) and theta (4–8 Hz) frequency bands (e.g., Andrillon et al., 2021; Hawes et al., 2020; Krueger et al., 2019; Nir et al., 2017; Pigarev et al., 1997; Poudel et al., 2021; Rector et al., 2005, 2009; Vyazovskiy et al., 2011). SLSW are generally spatially confined, primarily to frontal regions, and differ from the more distributed, global, and high amplitude slow waves characteristic of NREM sleep (D’Ambrosio et al., 2019; Krueger et al., 2019; Rector et al., 2009; Vyazovskiy et al., 2008). Importantly for PD, which is characterized by chronic neuroinflammation (Hirsch et al., 2012; Wang et al., 2015), animal studies have demonstrated a causal relationship between the release of pro-inflammatory factors (e.g., tumor necrosis factor alpha (TNFα); interleukins) and the local occurrence of SLSW (Krueger et al., 2011). Intracranial and cortical recordings in humans and animals indicate that, even during wakefulness, SLSW coincide with reduced neuronal firing patterns similar to sleep. Furthermore, these diurnal sleep intrusions are associated with several behavioral changes, including increased perceptual errors (Andrillon et al., 2021; Nir et al., 2017; Vyazovskiy et al., 2011), diminished vigilance (Andrillon et al., 2021), and transient cognitive slowing (Sheybani et al., 2023).

In addition to cognitive changes, diurnal sleep intrusions in awake individuals have been suggested to be associated with another dominant and clinically relevant non-motor symptom in PD: hallucinations (e.g., Arnulf et al., 2000; Comella et al., 1993; Kulisevsky & Roldan, 2004; Nomura et al., 2003; Manni et al., 2002). Hallucinations in PD consist of minor hallucinations (presence hallucinations, passage hallucinations, visual illusions) and complex visual hallucinations (for reviews, see Ffytche et al., 2017; Pagonabarraga et al., 2024), with minor hallucinations (Pagonabarraga et al., 2016) generally preceding complex visual hallucinations in the course of the disease (Goetz et al., 2006; Ffytche et al., 2017). Hallucinations have been linked with an increased cortical alpha-synuclein burden (Halliday & McCann, 2008; Harding et al., 2002; Onofrj & Gilbert, 2018; Papapetropoulos, 2006), alterations in brainstem and thalamic regions (e.g., Goetz et al., 2014; Hall et al., 2019; Ignatavicius et al., 2024, 2025; Thomas et al., 2023; Zarkali et al., 2022), as well as cortical regions (e.g., Bejr-Kasem et al., 2021; Bernasconi et al., 2021; Shine et al., 2015; Zarakali et al., 2022; for reviews, see Ffytche et al., 2017; Pagonabarraga et al., 2024). Minor and complex visual hallucinations have been linked to cognitive impairment and decline, suggesting a more malignant form of PD, advancing more rapidly and also involving cortical networks (Aarsland et al., 2000; Bejr-Kasem et al., 2021; Bernasconi et al., 2021, 2023; Diederich et al., 2009; Gryc et al., 2020). A separate line of work linked hallucinations to sleep disturbances (e.g., Goetz et al., 2008; Lenka et al., 2016; Onofrj et al., 2006) and it has been suggested that hallucinations in PD may either reflect daytime REM (Arnulf et al., 2000; Comella et al., 1993; Kulisevsky & Roldan, 2004; Nomura et al., 2003) or NREM intrusions into wakefulness (Manni et al., 2002). However, it is not known whether local sleep mechanisms during wakefulness, such as SLSW, are altered in PD and whether they are associated with a more malignant form of PD, characterized by hallucinations and cognitive impairment.

Here, we leveraged electroencephalography (EEG) resting-state recordings and in-depth neuropsychological evaluation and neuropsychiatric interviews conducted in a large sample of 84 patients with PD without dementia, with and without minor hallucinations, as well as in 30 healthy elderly control participants, to explore alterations in SLSW in PD and the hallucinatory trait. Hence, we quantified whether SLSW are altered as part of the neural degeneration occurring in PD compared to normal aging in healthy elderly. Furthermore, we determined whether SLSW are associated with minor hallucinatory traits, hallucinatory burden, and cognitive impairment. Our results provide novel and robust evidence that diurnal SLSW mediate perceptual disturbances and cognitive impairment in PD.

## Results

### Hallucinations (semi-structured interview)

Based on the *Hallucinations and Psychosis* item of the Movement Disorder Society Unified Parkinson’s Disease Rating Scale (MDS-UPDRS) Part I, item 1.2 (Goetz et al., 2008), and the administration of the semi-structured interview for the identification and characterization of psychotic phenomena in PD, 84 patients with PD were grouped into those who reported hallucinations (PD-H; N = 37) and those without any hallucination (PD-nH; N = 47). In the PD- H subgroup, 23 patients reported presence hallucinations, 19 patients reported passage hallucinations, and 16 patients reported visual illusions/pareidolia. In addition to those minor hallucinations, two patients reported complex visual hallucinations, two patients reported auditory hallucinations, three reported olfactory hallucinations, and none reported tactile hallucinations. It is important to note that a patient can experience more than one hallucination as part of the disease. Healthy elderly participants (HC; N = 30) reported no hallucinations.

### Demographical, clinical, and neuropsychological data

Across the three subgroups (PD-H; PD-nH, HC), our demographic data did not show any difference for age, sex, education, or depression (all p-values > 0.05; Table S1). There was a difference between the three groups for cognitive functions (Parkinson’s Disease-Cognitive Rating Scale (PD-CRS)), clinical Rapid eye movement Sleep Behaviour Disorder (RBD), and anxiety (Table S1). When comparing the two sub-groups of patients (PD-H vs. PD-nH), however, we found no differences; thus, the two patient subgroups did not significantly differ on most of the clinical demographic variables, except for daily levodopa equivalence (Table S1). Of note, PD-H reported significantly more (p-value < 0.001) subjective diurnal sleepiness (item from the MDS-UPDRS Part I, item 1.8 – Goetz et al., 2008) than PD-nH, but there was no significant difference in clinical RBD between PD subgroups (p-value = 0.075; Postuma et al., 2012). Furthermore, the increased somnolence in PD-H patients was confirmed when additionally controlling for RBD score. While earlier research associated excessive daytime somnolence to hallucinations in more advanced forms of PD (i.e., including those with dementia and complex visual hallucinations; Archibald et al., 2011), our findings reveal that this relationship emerges already at an earlier disease stage — especially in PD patients who only have minor hallucinations without any signs of dementia. Furthermore, our findings indicate that the increased daytime sleepiness observed in hallucinating PD patients is independent of dopaminergic medication and RBD. We also observed that both PD subgroups have more diurnal sleepiness than HC. This might be due to the known reduced sleep quality in PD or deficient arousal systems (Iranzo et al., 2024). Overall, our results suggest that subjective diurnal sleepiness is increased in PD, and even further in patients with minor hallucinations, extending previous findings (e.g., Fénelon et al., 2000).

### Sleep-like slow waves (SLSW)

We investigated several electrophysiological aspects of SLSW: first, the presence of SLSW, which can be observed in the delta (0.5-4Hz) and theta (4-8Hz) frequency bands (for a review, see Andrillon & Oudiette, 2023); second, the differences in SLSW features (density, peak-to-peak amplitude, and their cumulative sum) between the three subgroups; third, the associations between SLSW features and clinical variables, such as hallucinatory burden and cognitive impairment.

### Fronto-central theta SLSW density is enhanced in PD

First, we investigated whether the density (the average number of slow waves per minute over the whole recording, indicating the instant measure of events) of SLSW differs between the three sub-groups. We found a significant (p-values < 0.05; FDR-corrected; main effect of sub- groups; Figure 1A; Table S2) modulation of the density for theta SLSW. This significant modulation was localized over fronto-central electrodes (Figure 1A). Post-hoc analyses within the significant cluster (Figure 1A) showed that theta density is higher in PD-H vs. HC (p-values < 0.05, FDR-corrected; Table S3), and higher in PD-nH vs. HC (p-values < 0.05, FDR-corrected; Table S3), with no statistical density difference between PD-H vs. PD-nH (permutation p-values > 0.05, FDR-corrected; Table S3). No significant modulation of the density was found for the delta SLSW (no significant main effect of sub-groups, all p-values > 0.05; FDR-corrected; Figure 1B; Table S4).

**Figure 1.**
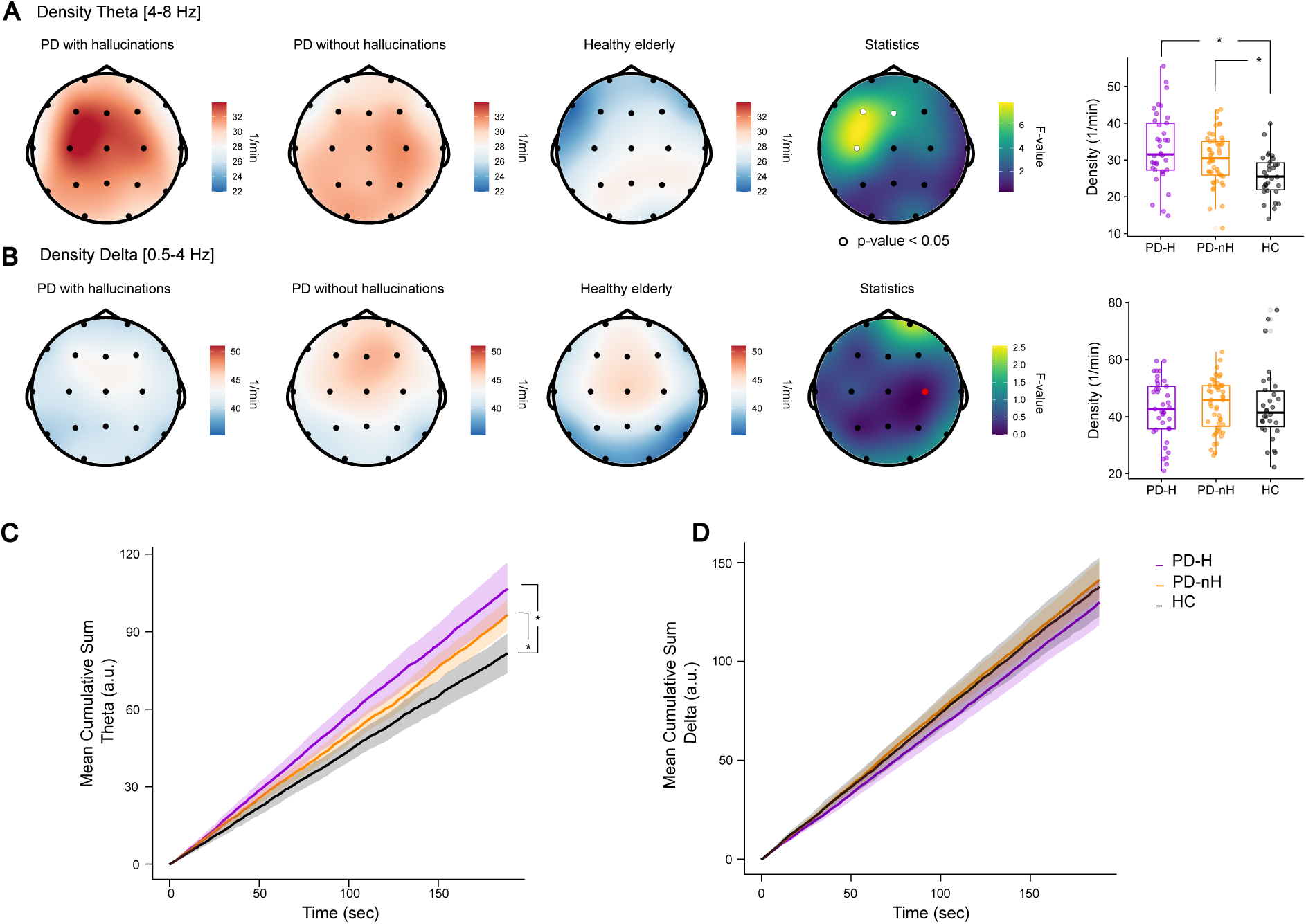
Fronto-central theta SLSW density is enhanced in PD. **A.** EEG topographies indicating the average theta SLSW density obtained for PD-H (left), PD-nH (center), and HC (right). The rightmost topography indicates the F-values of the one-way interaction between the three groups. Electrodes highlighted in white indicate significant interactions (p-values < 0.05, FDR-corrected). The box plots show the average density within the significant cluster and the post-hoc results. Single dots represent single individuals. **B.** EEG topographies indicating the average delta SLSW density obtained for PD-H (left), PD-nH (center), and HC (right). The rightmost topography indicates the F-values of the one-way interaction between the three groups; no significant electrode was observed after FDR correction. The box plot shows the density for the electrode C4 (highlighted in red; selected for visualization purposes only). For both subplots, each dot represents the average of a single participant. **C.** Mean cumulative sum of theta SLSW, on electrode C3. **D.** Mean cumulative sum of delta SLSW, on electrode C3.

Recent data (e.g., Boutin & Doyon, 2020; Champetier et al., 2023) showed that during sleep, specific sleep oscillatory activity can occur in clusters (“trains” or bursts). Therefore, we investigated whether this also applies to SLSW during wakefulness. That is, we assessed whether SLSW occurred homogenously or in bursts. Hence, we calculated the Fano Factor (FF; defined as the ratio of the variance to the mean number of slow waves occurring in a given time, providing a measure of variability in slow waves occurrence). Results show that the FF is close to 1, suggesting that SLSW occurrence follows a Poisson process rather than occurring in bursts. Moreover, this was the case for both the delta and theta SLSW (Table S5). We further investigated alterations in the SLSW temporal dynamics by computing the cumulative sum of SLSW events in time. The results corroborate the constant rate of SLSW events in PD and HC, with a higher accumulation of SLSW in PD (Figure 1C-D; Figure S1). Collectively, these results show that patients with PD (independent from the hallucinatory trait) have a higher theta (but not delta) SLSW density compared to HC, suggesting that parkinsonian neurodegeneration exacerbates theta SLSW density. Furthermore, these results also suggest that the SLSW event rate is constant over time, rather than occurring in clusters.

### Theta and delta SLSW have a higher peak-to-peak amplitude in PD-H

Next, we investigated whether the peak-to-peak amplitude of SLSW (Figure 2A) is modulated across the three subgroups and found a significant (p-values < 0.05; FDR-corrected; Table S6) modulation of the peak-to-peak amplitude across the three subgroups for both theta and delta SLSW (Figure 2B-C). That is, we found a significant (p-values < 0.05; FDR-corrected) modulation of the peak-to-peak amplitude across the three subgroups for the theta SLSW over fronto-parietal electrodes (Figure 2C), and post-hoc analyses showed that the peak-to-peak amplitude of SLSW is significantly higher in PD-H vs. PD-nH (Table S7). Furthermore, we observed that the peak-to-peak is significantly higher (p-values < 0.05; FDR-corrected; Table S7) in PD-H vs. HC, with no statistical difference (p-values > 0.05; FDR-corrected; Table S7) between PD-nH and HC (Figure 2B). Results for the delta SLSW were similar, for which the significant (p-values < 0.05; FDR-corrected; Table S8) peak-to-peak amplitude modulation was localized over fronto-parietal electrodes (Figure 2C). Post-hoc analyses of this cluster of electrodes show that the peak-to-peak amplitude is significantly higher in PD-H vs. PD-nH (Figure 2C; Table S9). Furthermore, our results show that the peak-to-peak of SLSW is significantly higher in PD-H vs. HC (p-values < 0.05; FDR-corrected; Table S9) and that there was no statistical difference (p-values > 0.05; FDR-corrected; Table S9) when comparing PD- nH to HC (Figure 2C). These results show that the peak-to-peak amplitude of delta and theta SLSW are modulated by the hallucinatory trait, being the largest in PD-H (for SLSW downward and upward slopes see Figures S2-S3 and Tables S10-S17).

**Figure 2.**
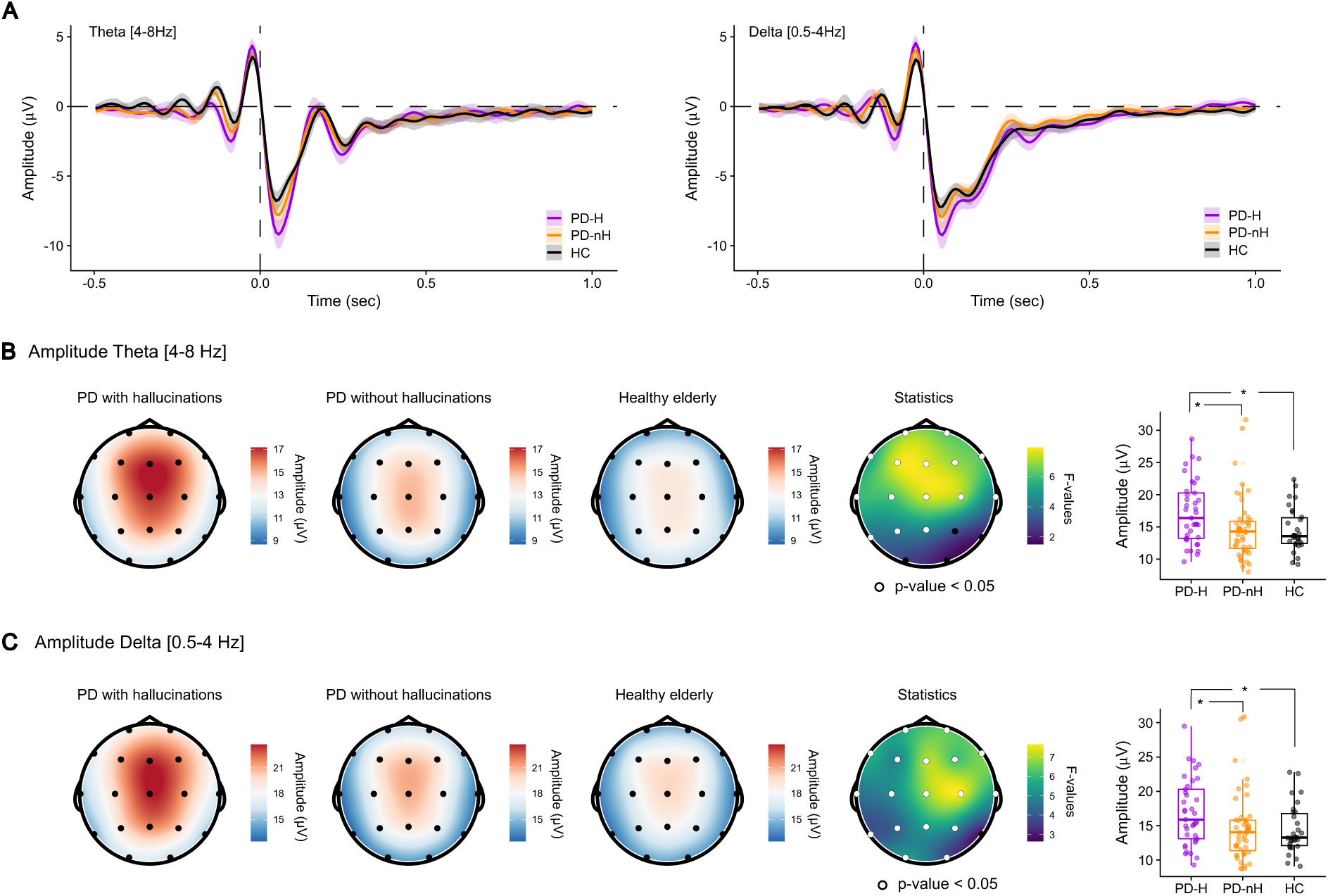
SLSW peak-to-peak amplitude is higher in PD-H than in PD-nH and HC. **A.** Average waveform of the SLSW detected over electrode Fz during the resting-state period. The rightmost plot shows the theta SLSW (4- 8Hz), PD-MH (purple), PD-nH (orange), HC (black). The thicker line represents the mean of the group; the shaded area represents the 95% confidence interval. Left plot shows delta SLSW (0.5-4Hz), PD-MH (purple), PD-nH (orange), HC (black). The thicker line represents the mean of the group; the gray shaded area represents the 95% confidence interval. **B.** EEG topographies indicating the average across subjects’ slow-waves peak-to-peak amplitude obtained for the theta SLSW for PD-H (left), PD-nH (center), and HC (right). The rightmost topography indicates the F-values of the one-way interaction between the three groups (white dots indicate p-values < 0.05, FDR-corrected). The box plots indicate post-hoc results analyses of the significant fronto-central cluster; each dot represents the average of a single participant. **C.** EEG topographies indicating the average across subjects for the delta SLSW peak-to-peak amplitude obtained for PD-H (left), PD-nH (center), and HC (right). The right topography indicates the F-values of the one-way interaction between the three groups (white dots indicate p-values < 0.05, FDR-corrected). The box plots indicate post-hoc results analyses of the significant fronto-central cluster; each dot represents the average of a single participant.

### The cumulative sum of peak-to-peak amplitude of theta and delta SLSW as an index of PD and PD hallucinations

To further characterize the alterations in the SLSW in PD and PD with hallucinations, we computed the cumulative sum of the peak-to-peak amplitude. This metric provides an index of SLSW alterations, capturing intensity, frequency, and temporal dynamics all in one. Our results show alterations for both the theta and delta SLSW, as well as a (partially) distinct pattern for theta and delta SLSW (Figure 3). For the theta SLSW cumulative sum of peak-to-peak amplitude, we found a significant (p-values < 0.05; FDR-corrected; Table S18) modulation across the three subgroups, localized over fronto-central electrodes (Figure 3A). Post-hoc analyses showed that the slope of the cumulative sum is significantly steeper in PD- H vs. PD-nH and in PD-H vs. HC (Table S19). Furthermore, we also observed that the slope of accumulation is significantly steeper in PD-nH vs. HC (Figure 3A; Table S19). Results for the delta SLSW showed a different pattern compared to theta SLSW (Figure 3B). With a difference in slope more localized over posterior-central electrodes, the slope of accumulation is steeper in PD (both PD-H and PD-nH) than in HC (Figure 3B; Table S20). Post-hoc analyses showed that the slope of the cumulative sum is significantly steeper in PD-H vs. PD-nH and in PD-H vs. HC (Table S19). Furthermore, we also observed that the slope of accumulation is significantly steeper in PD-nH vs. HC (Figure 3A; Table S19). Post-hoc analyses showed that the slope of the cumulative sum is significantly steeper in PD-H vs. PD-nH and in PD-H vs. HC (Table S21). Furthermore, we did not observe a significant difference in slope of accumulation between PD-nH and HC (Figure 3A; Table S21).

**Figure 3.**
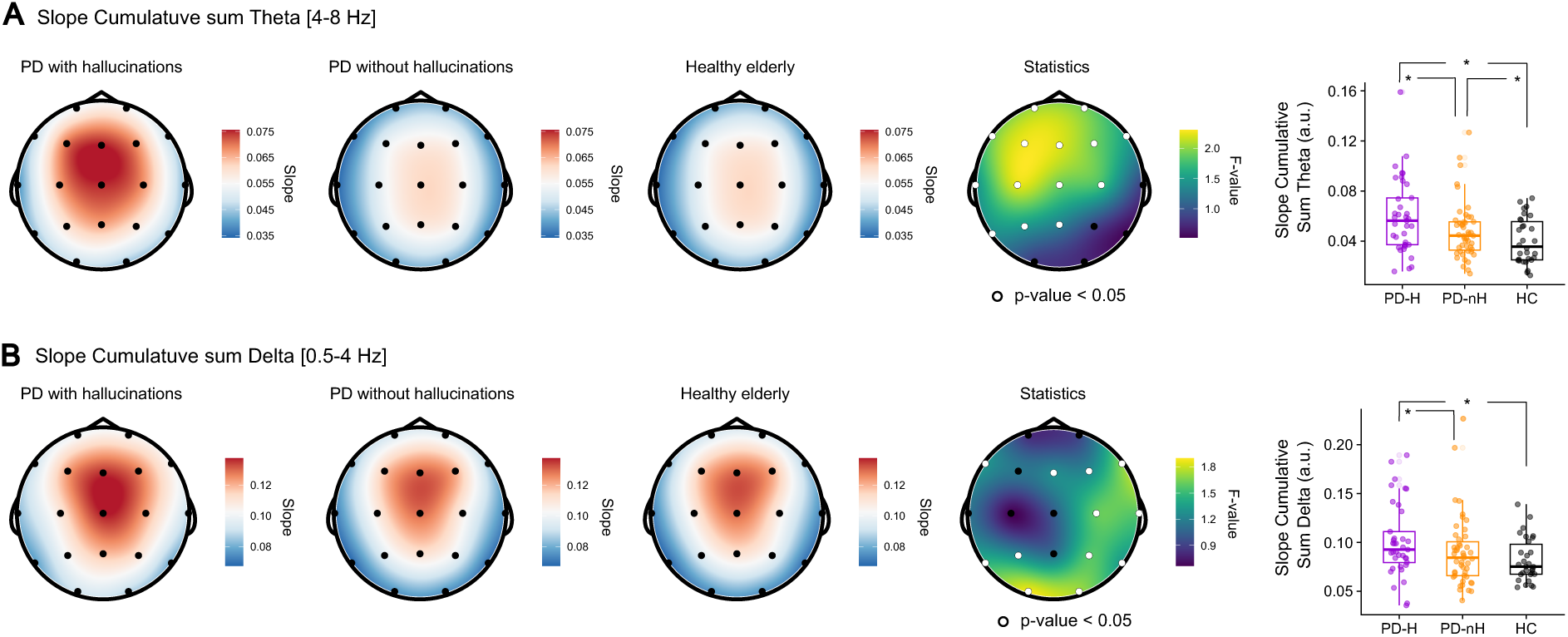
SLSW cumulative sum of the peak-to-peak amplitude is modulated by PD and by the hallucinatory trait. **A.** EEG topographies indicating the average across subjects of the slopes of the accumulation rate of theta SLSW obtained for PD-H (left), PD-nH (center), and HC (right). The rightmost topography indicates the F-values of the one-way interaction between the three groups (white dots indicate significant electrodes with permutation p- values < 0.05, FDR-corrected). The box plots show the average density within the significant cluster and the post-hoc results. Single dots represent a single individual. **B.** EEG topographies indicating the average delta SLSW slopes of the accumulation rate obtained for PD-H (left), PD-nH (center), and HC (right). The rightmost topography indicates the F-values of the one-way interaction between the three groups (white dots indicate significant electrodes with permutation p-values < 0.05, FDR-corrected). The box plots show the average density within the significant cluster and the post-hoc results. Single dots represent a single individual.

### Theta SLSW peak-to-peak amplitude uncovers a more severe hallucinatory burden

We explored whether SLSW peak-to-peak amplitude (on each electrode independently) is associated with the hallucinatory burden (i.e., sum between the count of different hallucinations experienced by patients and the score of the MDS-UPDRS, Part I, item 1.2, assessing hallucinations and psychosis severity). Our results show that a higher peak-to-peak amplitude for the theta SLSW are associated with a higher hallucinatory burden, observed over fronto-central regions (p-values < 0.05; FDR-corrected; Figure 4; Table S18; for delta SLSW, similar trends of significance were observed but did not survive correction for multiple comparisons, Table S23). Similar results were observed when we used the cumulative sum of peak-to-peak amplitude (Tables S24-25). A higher number of different minor hallucinations has been associated a higher prevalence of complex visual hallucinations, suggesting that minor hallucinations might have a predictive role of more malignant forms of PD (Zhang et al., 2025). Our results further strengthen the link between minor hallucinations and a malignant form of the disease, by showing the association between theta SLSW and the hallucinatory burden.

**Figure 4.**
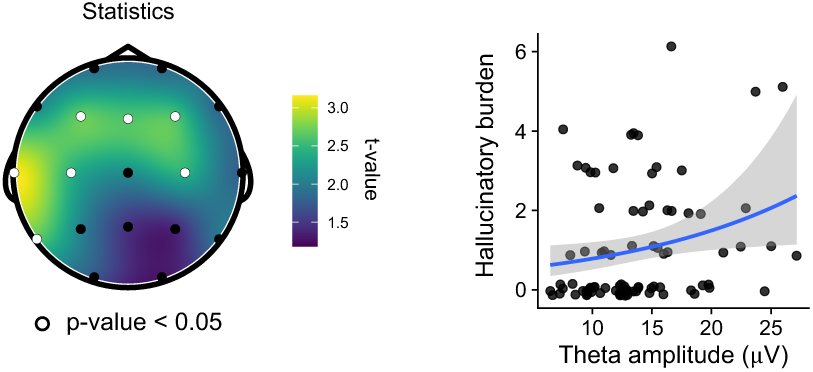
Increased SLSW amplitude is associated with a more severe hallucinatory burden. Higher SLSW theta peak-to-peak amplitude is associated with a more severe hallucinatory burden. The topography shows the t-values obtained from a quasi-Poisson ANCOVA, conducted to model the effect of the peak-to-peak amplitude (fixed effect) on the hallucination burden (dependent variable). The model included age, sex, and years of education as covariates of no interest (one fit per electrode); the white electrodes correspond to a significant (p-values < 0.05; FDR-corrected) prediction. In the scatter plot, each dot represents the average of the significant cluster for each patient. The right plot shows the result for electrode F3, which is part of the significant cluster (white electrodes on the topography).

### Prominent theta SLSW density and peak-to-peak amplitude are associated with more severe cognitive impairment

Based on prior evidence that (reduced) fronto-central slow waves during sleep are associated with cognitive impairment in neurodegenerative diseases (Schreiner et al., 2021; Sharon et al., 2025), here, we analyzed (on each electrode) whether the density and/or amplitude of SLSW during wakefulness are associated with cognitive impairment. Our results show that, independently of the subgroups, more severe cognitive impairment (measured with the PD-CRS total score) is associated with higher fronto-parietal theta SLSW density (p-values < 0.05; FDR-corrected; Figure 5A; Table S27). This association was not observed for the delta SLSW density (Table S28). Furthermore, we observed that the peak-to-peak amplitude is also associated with cognitive impairments for the theta SLSW (Figure 5B; p-values < 0.05; FDR-corrected; Table S29). This was not the case for the delta SLSW peak-to-peak amplitude (Table S30). Our results showed that SLSW are neither associated with diurnal somnolence nor the presence of RBD (Tables S31-S34).

**Figure 5.**
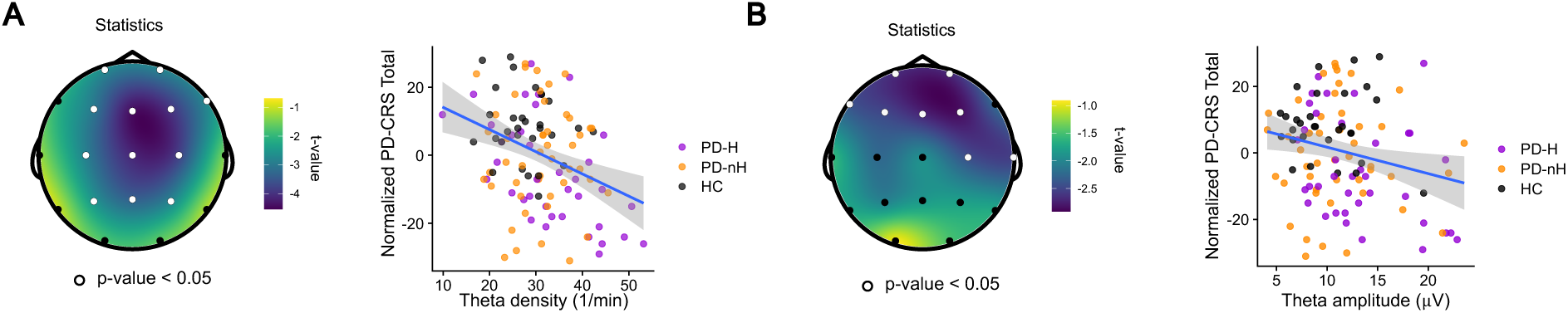
Increased SLSW uncovers more severe cognitive impairment. **A.** Higher SLSW theta is associated with more severe cognitive impairment. The topography shows the t-values of the linear regressions (one per electrode); the white electrodes correspond to a significant (permutation p-values < 0.05; FDR-corrected) association. In the scatter plot, each dot represents one individual, and the density on the electrode Cz (part of the significant cluster). **B.** The severity of the cognitive impairment is also associated with the theta peak-to-peak amplitude. In the scatter plot, each dot represents the one individual, and the density on the electrode Fp2 (part of the significant cluster).

## Discussion

Growing evidence suggests that there are periods of brain activity in fully awake healthy individuals that resemble NREM sleep slow waves, even though such SLSW are more focal and frontally localized. In the present study, we conducted resting-state EEG and in-depth psychiatric and neuropsychological assessments in 84 non-demented PD patients and 30 healthy elderly participants, to determine the presence of altered SLSW in PD and their role in hallucinations and cognitive impairment. First, we report that PD patients have higher diurnal sleepiness than healthy elderly, especially patients with minor hallucinations. Second, we discover that there is an increased density of SLSW in PD when compared to elderly HC (Figure 1). Third, we show that the hallucinatory trait in PD, indicative of a more malignant form of PD, is characterized by an even stronger alteration of SLSW (compared to PD patients without any hallucinations), exhibiting higher SLSW amplitude and accumulation over fronto- parietal regions (Figures 2 and 3). Finally, we demonstrate that the degree of SLSW alterations predicts the hallucinatory burden (Figure 4) as well as cognitive impairment (Figure 5). Collectively, these results provide novel and robust insights into the implications of SLSW in PD and their role in important non-motor symptoms: aberrant perceptions and cognition. While slow waves during sleep have positive physiological functions, related to homeostasis and metabolic waste clearance (e.g., reducing α-synuclein burden), exacerbated SLSW during the day may represent reactions related to the PD neuropathology, at the cost of more hallucinations and cognitive impairment.

Global slow waves occurring during NREM sleep have been extensively investigated in neurodegenerative disease (e.g., Ju et al., 2014, 2017; Lee et al., 2020b; Morawska et al., 2021), revealing an important role of such slow waves in neural homeostasis and metabolic waste clearance (e.g., α-synuclein and beta-amyloid reduction). Growing evidence also showed intrusion of sleep-like cortical dynamics during wakefulness, yet more localized to frontal regions. These global (sleep-related) and local (during wakefulness) transitions toward sleep are characterized by the same neural signature: the occurrence of high-amplitude slow waves. While previous studies have established that PD patients have reduced NREM global slow waves during sleep (e.g., Chen et al., 2023), the role of local slow waves (SLSW) has not been explored in PD. Our results show another face of PD by highlighting the opposite pattern for SLSW during wakefulness. We demonstrate that patients with PD — regardless of their hallucinatory trait — exhibit a higher density and a more rapid accumulation of SLSW compared to healthy elderly. The observed increase in SLSW density might be driven by the neuroinflammation associated with PD, as an increase in pro-inflammatory factors such as interleukins and TNFα have been associated with a local increase in SLSW in rodents (Hirsch et al., 2012; Wang et al., 2015). The increase in SLSW could also reflect a compensatory mechanism to the underlying α-synuclein pathology, by promoting the metabolic waste clearance during wakefulness (Yang et al., 2024). Increased SLSW has been associated with higher sleep pressure, longer time spent awake (e.g., Bernardi et al., 2015; Hung et al., 2013). Although somnolence might partially explain the increased SLSW that we observed in PD; we did not find any significant association between somnolence scores and SLSW density (nor the presence of clinical RBD).

Critically, the present data show that the increase in SLSW (density and amplitude) is especially strong in PD patients with a psychiatric non-motor symptom: minor hallucinations. SLSW represent a state during which cortical neurons briefly go “silent” (Bernardi et al., 2015; Nir et al., 2011; Vyazovskiy et al., 2011), leading to impaired information processing and impaired performance in several behavioral tasks (Andrillon et al., 2019; Bernardi et al., 2015; Hung et al., 2013; Nir et al., 2017; Vyazovskiy et al., 2011). Moreover, the presence of SLSW (in healthy individuals) has been linked to fluctuations in levels of attention (Andrillon et al., 2021). Based on the present data, we argue that the increased neural silencing caused by increased SLSW activity in PD facilitates the occurrence of hallucinations through short but repeated periods characterized by fluctuating vigilance and impaired attentional mechanisms. Alterations in attentional mechanisms have previously been shown to facilitate hallucinations in PD (Ignatavicius et al., 2025; Manni et al., 2011; Pagonabarraga et al., 2024; Shine et al., 2014). It was shown that a stronger segregation of different attentional regions is observed in PD patients with visual complex hallucinations (Zarkali et al., 2022). Periods with fluctuating vigilance, characterized by a more prominent occurrence of SLSW, have been associated with increased mind-wandering (Andrillon et al., 2021; Philippi et al., 2021), leading to a disconnection from the environment and an over reliance on internal cognitive mechanisms (e.g., perceptual experiences) (Goupil & Bekinschtein, 2012; Kam et al., 2011; Tagliazucchi & Laufs, 2014). Some have suggested that hallucinations may be considered spontaneous intrusions into the mind of dreams/mental images, possibly occurring during fluctuations between wakefulness and REM/NREM sleep (Arnulf et al., 2000; Kulisevsky & Roldan, 2004; Manni et al., 2002; Shine et al., 2015). Hence, during such periods of reduced vigilance, PD with hallucinations may experience increased access to internally generated mental states (i.e., mental perceptions and/or imagery in the absence of external stimuli) (Shine et al., 2015), which can be perceived as vivid and externally generated stimuli (Dijkstra et al., 2021, 2025; Dijkstra & Fleming, 2023; Shine et al., 2015; Walpola et al., 2020; Zeman, 2024; Zeman et al., 2015, 2020).

Several PD pathomechanisms might underlie the altered SLSW we observed in PD patients with hallucinations. First, associations between the density and accumulation of α-synuclein in subcortical and cortical regions and (complex visual) hallucinations in PD (Halliday & McCann, 2008; Jacobson et al., 2014; Onofrj et al., 2006, 2007; Onofrj & Gilbert, 2018; Papapetropoulos, 2006). Accordingly, the increased SLSW we observed in PD with hallucinations might be a consequence of the α-synuclein burden and an increased attempt to clear it. Second, preliminary evidence suggests that neuroinflammation, which leads to increased SLSW (Wang et al., 2015), might also be related to hallucinations (for a review, see Swann et al., 2024). A third plausible explanation for the increased SLSW observed in PD, especially in those with hallucinations, might be thalamocortical dysrhythmia (Collerton et al., 2023), which has been associated not only with the presence but also with the severity of hallucinations in PD (e.g., Goetz et al., 2014; Hall et al., 2019; Ignatavicius et al., 2024, 2025; Thomas et al., 2023; Zarkali et al., 2022). It has been proposed that a “bottom-up” synchronization mechanism, mediated by subcortical regions (including the brainstem and thalamus), results in cortical slow waves, particularly in frontomedial and sensorimotor areas (Siclari et al., 2014; Stephan et al., 2025). Accordingly, SLSW might be interpreted as sleep intrusions during wakefulness, with neurophysiological changes at the frontal level caused by the disrupted functionality of brainstem and thalamic structures. Future research should examine whether increased SLSW results from one or several of these pathologies.

Concerning cognitive function, previous findings have shown that during sleep, reduced NREM slow waves are associated with more severe cognitive impairment in PD (Schreiner et al., 2021). Enhanced intrusions of SLSW during wakefulness have been shown to have detrimental effects on cognitive performance, leading, for example, to slower reaction times (Andrillon et al., 2021; Sheybani et al., 2023) as well as increased errors in cognitive paradigms (e.g., Andrillon et al., 2021; Nir et al., 2017). Our results show that more prominent SLSW are associated with a more severe impairment in global cognitive functions, which have been assessed with in-depth neuropsychological evaluations. This association was found in both tested populations, PD and healthy elderly. The negative impact of SLSW on cognition might be due to the neural silencing that is typically associated with slow waves, especially with the negative deflection of SLSW. Hence, increased SLSW, which might be due to neuroinflammation, neural degeneration, and/or an increased attempt of α-synuclein clearance, indicates prolonged periods of a neural silent off-period (or transition between on and off), which have a detrimental impact on cognition. Both neuroinflammation and α-synuclein are associated with a higher risk of dementia (Aarsland et al., 2005; Kouli et al., 2024). Prominent slow waves (especially theta) in PD and dementia with Lewy bodies have been associated with cognitive decline (e.g., Babiloni et al., 2020; Bernasconi et al., 2023; Bosboom et al., 2006; Hassan et al., 2017; Onofrj et al., 2019) corroborating the relevance of those slow waves for cognitive alterations.

In conclusion, our study provides novel evidence on the adverse role of SLSW in PD, the hallucinatory trait, and cognitive functions. SLSW could be a new prodromal marker of PD and might be especially useful for identifying individuals at risk of more malignant forms of the disease, which are characterized by non-motor symptoms such as psychosis and dementia. Furthermore, treatments addressing disturbances in slow waves could represent an interesting new therapeutic approach aimed at diminishing these highly clinically relevant non-motor symptoms and tackling the impairments engendered by cognitive decline in PD.

## Limitations of the study

Our findings should be considered in the context of the following limitations. First, although the use of an EEG system with a limited number of electrodes has several advantages for clinical use and patient comfort, a higher number of electrodes will allow for more extensive analyses (for example, source localization). Second, while our results show the relevance of SLSW for hallucinatory traits, future studies should investigate the role of SLSW for hallucinatory states. That is, the role of SLSW should be investigated while clinically relevant hallucinations are induced experimentally (Bernasconi et al., 2021; 2022; Albert et al., 2024). Third, future experiments should investigate the direct association between SLSW, mind-wandering, and hallucinations. Fourth, hallucinations were evaluated using the *Hallucinations and Psychosis* item of the MDS-UPDRS Part I and a semi-structured interview addressing various types of hallucinations associated with PD. In future studies, additional screening questionnaires should be used to explore further aspects of hallucinations, such as frequency and insight.

## Materials and methods

### Participants

One hundred and four individuals participated in the current study. Eighty-four individuals were diagnosed with PD, and thirty were healthy elderly (HC). All those fulfilling MDS new criteria for PD with minor hallucinations (PD-H) — sense of presence, passage hallucinations, visual illusions and/or pareidolias, but also complex visual hallucinations, tactile, and olfactory hallucinations (N = 37) — and without any hallucinations (PD-nH; N = 47) were prospectively recruited from a sample of outpatients regularly attending to the Movement Disorders Clinic at Hospital de la Santa Creu i Sant Pau, Barcelona. Individuals were diagnosed with PD by a neurologist with expertise in movement disorders. Each individual was interviewed regarding disease onset, medication history, current medications, and dosage (levodopa daily dose and dopaminergic agonist-equivalent daily dose). Motor status and stage of illness were assessed by the MDS-UPDRS Part III scale (Goetz et al., 2008). The two subgroups of patients were comparable in sex, age, disease duration, dopaminergic doses, dopaminergic agonists, motor severity, sleep disturbances, and cognition.

Exclusion criteria included a history of major psychiatric disorders, cerebrovascular disease, and conditions known to impair mental status other than PD. Patients with focal abnormalities in MRI or non-compensated systemic diseases (i.e., diabetes, hypertension) were also excluded. In patients with motor fluctuations, cognition was examined during the best “on” state. All participants were on stable doses of dopaminergic drugs during the 4 weeks before inclusion. Patients were included if the hallucinations remained stable during the 3 months before inclusion in the study. No participant had used or was using antipsychotic medication. Participants who at baseline scored below the cut-off score for dementia — indicated by a score of 63 points on the Parkinson’s Disease-Cognitive Rating Scale (PD-CRS) (Emre et al., 2007) — and/or a Montreal Cognitive Assessment (MoCA) score < 18 were excluded from the study (Nasreddine et al., 2005). Clinical RBD was assessed with the Single-Question Screen for REM Sleep Behavior Disorder (RBD1Q) (Postuma et al., 2012). To quantify the level of anxiety and depression in patients, we used the Hospital Anxiety and Depression Scale (HADS) (Stern, 2014), and to quantify the level of apathy, we used the Starkstein Apathy Scale (Starkstein et al., 1992).

Informed consent to participate in the study was obtained from all participants according to the Declaration of Helsinki. The study was approved by the local ethics committee (#IIBSP-PAR-2019-17).

### Hallucinations and cognitive functions assessments

Presence and type of minor hallucinations was assessed using the *Hallucinations and Psychosis* item of the MDS-UPDRS Part I (Goetz et al., 2008) and a semi-structured interview covering the different types of minor hallucinations associated with PD. The *Hallucinations and Psychosis* item is a clinical scale, rated on a 0 to 4 spectrum: 0 corresponds to absence of hallucinations; 1, Slight: Illusions or non-formed hallucinations, but patient recognizes them without loss of insight; 2, Mild: Formed hallucinations independent of environmental stimuli. No loss of insight; 3, Moderate: formed hallucinations with loss of insight; and 4, Severe: Patient has delusions or paranoia. The semi-structured interview used in this study has been used and published in previous studies of our group (Pagonabarraga et al., 2016; see mds26432-sup-0001-suppappendix.doc; Bejr-Kasem et al., 2019; see mds27557-sup-0001-Supinfo.docx), and is used for the identification and characterization of different psychotic phenomena associated with PD. We categorize as minor hallucinators those participants with a score = 1 in the *Hallucinations and Psychosis* item of the MDS-UPDRS Part I, and reported a sense of presence, passage hallucinations, and/or visual illusions using the semi-structured interview, at least weekly during the last month. In summary, patients were included in the PD-MH (i.e., with minor hallucinations) group if they had hallucinations at least monthly, and if these phenomena were present during the three months before inclusion in the study. Cognition was assessed by the PD-CRS (Pagonabbarraga et al., 2008), which is a cognitive scale specifically designed to capture the whole spectrum of cognitive functions impaired over the course of PD (Pagonabarraga et al., 2008). This battery is composed by a total of 9 tasks explicitly designed for a brief and separate scoring procedure including frontal-subcortical tasks (sustained attention, working memory, alternating and action verbal fluency, clock drawing, immediate and delayed free recall verbal memory) as well as posterior cortical tasks (confrontation naming, clock copying). The frontal-subcortical assessment score ranges from 0 to 114 points, the posterior assessment score from 0 to 20 points. The sum of the two scores is added to give the total score of the PD-CRS (0-134).

### Resting-state EEG

Resting-state EEG data was collected using a 19-channel EEG system. Resting-state EEG data were collected with eyes open for a period lasting 5 minutes. Neuropsychiatric, neuropsychological and EEG data were collected during the best ON dopaminergic state for each patient. The EEG recordings were always performed between 9AM and 2PM.

### EEG acquisition and preprocessing

EEG acquisition was done using the Brain Vision Recorder and BrainAMP system (Brain Products GmbH; Germany). Data were analyzed off-line with the EEGLAB toolbox (version 2023.0; Delorme & Makeig, 2004; http:// sccn.ucsd.edu/wiki/EEGLAB) for MATLAB, and Fieldtrip toolbox (version -20240110; www.fieldtriptoolbox.org; Oostenveld et al., 2011).

Continuous EEG was acquired at 250 Hz from 19 standard scalp sites (Fp1-2, F3-4, C3-4, T3-4, T5-6, P3-4, O1-2, F7-8, Fz, Cz, Pz) using passive tin electrodes mounted in an elastic cap and referenced to the two mastoid leads. Vertical eye movements were monitored using a bipolar montage with two electrodes linked together and placed below each eye referenced to a third electrode placed centrally above the eyes. Horizontal eye movements were monitored using two electrodes placed on the external canthi of each eye. Electrode impedances were kept below 5 kOhm. The electrophysiological signals were filtered with a bandpass of 0.1–40 Hz with a Butterworth filter of 5^th^ order (using Fieldtrip). Line noise was removed with *pop_cleanline (EEGlab function)*. Bad channel detection (flatline criterion = 5s; channel criterion = 0.8) and artifact subspace reconstruction (burst criterion = 20) as implemented via the *clean_rawdata* plugin (Mullen et al., 2015).

We computed the independent component analysis (ICA) (Makeig et al., 1996) on a copy of the EEG data that were high-pass filtered at 1Hz. The number of independent components (ICs) computed corresponded to 99% of the variance. We used SASICA (Chaumon et al., 2015) with default parameters to automatically select ICs for rejection and visually inspected all components. ICs reflecting eye blinks, saccades, or noise indicated by SASICA (and visual inspection) were identified and removed. ICs weights kept were then backprojected to the original EEG data. Any channel rejected prior to the ICA was interpolated using spherical interpolation. To avoid confounds in the detection and analyses of slow waves (see below) due to different EEG data lengths among patients, all the data were trimmed to the shortest recording (3.14 minutes) observed among all patients.

### Detection of slow waves

The detection of sleep-like slow waves in wakefulness was based on previous algorithms devised to automatically detect slow waves during NREM sleep (e.g., Andrillon et al., 2021). The pre-processed EEG signal was down-sampled to 125 Hz and low-pass filtered at 10Hz band, with a Butterworth filter 2^nd^ order (using Fieldtrip version 20240110). All waves were detected by locating the negative peaks in the filtered signal. For each wave, the following parameters were extracted: start and end point (defined as zero-crossing, respectively, prior to the negative peak of the wave and following its positive peak), negative peak amplitude and position in time, positive peak amplitude and position in time, peak-to-peak amplitude, downward (from start to negative peak) and upward (from negative to positive peak) slopes.

To reduce the false detection of these artefacts as candidate slow waves, we excluded waves with a positive peak >75 µV. We also excluded waves within 1s of large-amplitude events (>150 µV of absolute amplitude). Finally, for delta slow waves we selected all waves that corresponded to a frequency within 0.5 and 4Hz, for the theta slow waves we selected all waves that corresponded to a frequency within 4 and 8Hz. To avoid introducing biases among subgroups, we did not apply any peak-to-peak amplitude threshold (e.g., top 10%) when selecting slow waves.

### Accumulation rate of SLSW

The accumulation rate was calculated first by calculating the cumulative sum of the occurrence of slow waves over time (using *cumsum* in Rstudio), for each electrode, subject, and frequency band separately. Then, to assess whether the accumulation rate was different among sub-groups, we fitted a linear regression on the accumulation rate (for each electrode, subject, and frequency band), extracted the slope, and assessed the statistical significance with a permutation test for ANOVA (10,000 iterations; permuco; Frossard & Renaud, 2021; R package), with slope as the dependent variable, sub-groups as the independent variable and age, sex, and years of education as covariates of no interest. Correction for multiple comparisons for the number of electrodes was obtained with False Discovery Rate (FDR) (Benjamini & Yekutieli, 2001).

### Statistical analysis

#### Clinical-demographic variables

Statistical difference between PD-MH, PD-nH, and HC on the measured clinical and demographic variables assessed using Welch test, Kruskal–Wallis test, Fisher Exact test, or Mann–Whitney U test, (see legend of Table S1 for details).

#### EEG sleep-like slow waves’ features analyses

All main statistical models compared the three groups (either with ANCOVA, linear regression, or linear mixed models depending on the data used as dependent variable), with age, sex, and education as covariates of no interest. Hence, to investigate significant modulations of the slow waves’ features among the sub-groups of participants we conducted permutation test for ANCOVA (10,000 iterations; permuco; Frossard & Renaud, 2021; R package)) for features that had only one value per electrode and subject (e.g., density and accumulation rate). For features that had multiple values per electrode and subject (i.e., peak-to-peak amplitude, up- and down-slope) we applied linear mixed-effects models (afex package; Singmann et al., 2024), with sub-groups of participants as fixed effects and a random intercept for each participant. Significance of fixed effects was estimated using Satterthwaite’s approximation for degrees of freedom of F statistics. For all models, age, sex, and years of education were used as covariates of no interest. Statistical models were applied independently to each electrode. Correction for multiple comparisons for the number of electrodes was obtained with FDR (Benjamini & Yekutieli, 2001). Contrast sum was used for the categorical variables in all the analyses performed. Contrast sum was used for the categorical variables in all the analyses performed.

#### EEG sleep-like slow waves’ features associated with clinical-demographic variables

To investigate significant associations between clinical variables and sleep-like slow waves we conduced linear regressions, with significance assessed with permutation test (10,000 iterations; permuco; Frossard & Renaud, 2021; R package). The statistical tests were applied to each electrode and frequency band separately. All models had the SLSW feature as the independent variable, and age, sex, and years of education as covariates of no interest. Correction for multiple comparisons for the number of electrodes was obtained with FDR (Benjamini & Yekutieli, 2001).

To investigate the association between SLSW and the hallucination burden a generalized ANCOVA was conducted using a generalized linear model (GLM) with a quasi-Poisson distribution. The dependent variable was the total number of hallucinations, and the primary predictor of interest was SLSW amplitude. Age, sex and years of education were included as covariates of no interest. Contrast sum was used for the categorical variables in all the analyses performed.

To investigate the association between SLSW and the hallucination burden, quasi-Poisson ANCOVA was conducted to model the effect of the peak-to-peak amplitude (fixed effect) on the hallucination burden (dependent variable). The model included age, sex, and years of education as covariates of no interest (one fit per electrode).

## Supporting information

Supplementary Results

## Acknowledgments

This research was supported by the Swiss National Science Foundation [grant n. 188798], CARIGEST SA (Fondazione Teofilo Rossi di Montelera e di Premuda and a second one wishing to remain anonymous) and Parkinson Suisse to O.B.; the Swiss National Science Foundation [grant n. 221182] and the Leenaards Foundation to F.B.; the Synapsis Foundation to O.B and F.B.; CIBERNED (Carlos III Institute) and FIS grant PI18/01717 to J.K.; Instituto de Salud Carlos III (ISCIII), Spain, to J.K.; PERIS, expedient number SLT008/18/00088 Generalitat de Catalunya to J.P.

## Code availability

Code will be made available via Gitlab (https://gitlab.epfl.ch/fbernasc/slsw_hallucinations.git) after publication acceptance.

## Data availability

Data will be made available via Gitlab (https://gitlab.epfl.ch/fbernasc/slsw_hallucinations.git) after publication acceptance.

## Authors contribution

Authors’ contributions according to the CRediT taxonomy (see http://credit.niso.org/).

Conceptualization: F.B., O.B.; additionally, T.A. and J.C.F. provided ideas and contributed to the evolution of the overarching research goals and aims.

Validation: F.B.

Formal analysis: F.B. and T.A. Investigation: J.P., J.K. Ressources: F.B., J.P., J.K., O.B.

Data curation: F.B., J.P., J.K., SMH Writing – Original Draft: F.B. and O.B. Writing – Review & Editing: all authors Visualization: F.B.

Project administration: F.B., O.B. Funding acquisition: O.B., F.B., J.K., J.P.

