## Supplementary Results for "Sleep-like slow waves during wakefulness uncover a malignant form of Parkinson’s disease"

#### *Demographical, clinical, and neuropsychological data*

| | PD-H (N = 37) | PD-nH (N = 47) | HC (N = 30) | *PD-H vs. PD-nH vs. HC (p-value)* | *PD-H vs. HC (p-value)* |
|---|---|---|---|---|---|
| Age (years) | 65.5 ± 8.4 | 65.5 ± 8.5 | 65.5 ± 8.6 | 0.0485 a | 0.125 b |
| Sex (male/female) | 28/9 | 28/19 | 17/13 | 0.194 c | 0.163 c |
| Education (years) | 12.7 ± 4.1 | 12.7 ± 4.2 | 12.7 ± 4.3 | 0.421 a | 0.774 b |
| Somnolence (MDS-UPDRS 1.8) | 1.1 ± 0.9 | 0.47 ± 0.7 | 0.23 ± 0.4 | <0.001 a | <0.001 b |
| PD-CRS total score | 89.9 ± 15.4 | 94.9 ± 15.5 | 103.7 ± 10.2 | <0.001 a | 0.112 b |
| MoCA score | 25.3 ± 3.4 | 25.7 ± 3.0 | 28.7 ± 1.5 | <0.001 a | 0.684 b |
| RBD1Q (present/absent) | 19/17 | 15/31 | 0/30 | <0.001 c | 0.075 c |
| Depression | 2.3 ± 2.2 | 2.5 ± 2.8 | 1.2 ± 2.1 | 0.123 a | 0.076 b |
| Anxiety | 3.8 ± 2.7 | 3.5 ± 2.5 | 2.1 ± 2.5 | 0.451 a | 0.693 b |
| Apathy | 5.1 ± 6.9 | 4.5 ± 5.7 | 1.0 ± 2.3 | 0.032 a | 0.544 b |
| Disease duration (years) | 6.6 ± 3.4 | 4.6 ± 2.8 | - | - | 0.007 b |
| Equivalent dopamine agonists (mg/day) | 157.1 ± 130.4 | 161.2 ± 102.1 | - | - | 0.686 b |
| LEDD (mg/day) | 691.8 ± 342.4 | 476.3 ± 269.9 | - | - | 0.003 b |
| MDS-UPDRS Part III (ON state) | 25.3 ± 7.7 | 25.3 ± 8 | - | - | 0.976 d |
| Hoen & Yahr stage | 2.2 ± 0.3 | 2.0 ± 0.3 | - | - | 0.012 d |

**Table S1.** Demographical, clinical, and neuropsychological variables. LEDD indicates levodopa equivalent daily dose. MDS-UPDRS Part III was measured during the ON dopaminergic medication state. *a* Kruskal–Wallis test; *b* Mann-Whitney U test; *c* Fisher exact test, and *d* Welch test.

*Theta SLSW density is increased in PD*

| chan | effect | F | p | pfdr | Band | chan | effect | F | p | pfdr | Band |
|---|---|---|---|---|---|---|---|---|---|---|---|
| C3 | Age | 0.05 | 0.833 | 0.998 | theta | C3 | Group | 7.385 | 0.001 | 0.013 | theta |
| C4 | Age | 0.011 | 0.923 | 0.998 | theta | C4 | Group | 3.35 | 0.035 | 0.082 | theta |
| Cz | Age | 0.194 | 0.654 | 0.998 | theta | Cz | Group | 3.212 | 0.046 | 0.09 | theta |
| F3 | Age | 0.359 | 0.563 | 0.998 | theta | F3 | Group | 7.407 | 0.001 | 0.013 | theta |
| F4 | Age | 0 | 0.998 | 0.998 | theta | F4 | Group | 3.893 | 0.022 | 0.062 | theta |
| F7 | Age | 0.586 | 0.444 | 0.998 | theta | F7 | Group | 4.661 | 0.012 | 0.056 | theta |
| F8 | Age | 0.019 | 0.888 | 0.998 | theta | F8 | Group | 1.88 | 0.155 | 0.213 | theta |
| Fp1 | Age | 0.207 | 0.647 | 0.998 | theta | Fp1 | Group | 3.224 | 0.052 | 0.09 | theta |
| Fp2 | Age | 0.04 | 0.847 | 0.998 | theta | Fp2 | Group | 3.877 | 0.023 | 0.062 | theta |
| Fz | Age | 0.009 | 0.919 | 0.998 | theta | Fz | Group | 5.707 | 0.005 | 0.033 | theta |
| O1 | Age | 0.044 | 0.835 | 0.998 | theta | O1 | Group | 1.286 | 0.276 | 0.308 | theta |
| O2 | Age | 0.117 | 0.738 | 0.998 | theta | O2 | Group | 3.084 | 0.049 | 0.09 | theta |
| P3 | Age | 0.053 | 0.813 | 0.998 | theta | P3 | Group | 1.924 | 0.157 | 0.213 | theta |
| P4 | Age | 0 | 0.997 | 0.998 | theta | P4 | Group | 1.655 | 0.196 | 0.249 | theta |
| P7 | Age | 0.287 | 0.594 | 0.998 | theta | P7 | Group | 2.186 | 0.118 | 0.187 | theta |
| P8 | Age | 0.023 | 0.878 | 0.998 | theta | P8 | Group | 0.762 | 0.47 | 0.47 | theta |
| Pz | Age | 0.074 | 0.786 | 0.998 | theta | Pz | Group | 1.564 | 0.212 | 0.252 | theta |
| T7 | Age | 0.048 | 0.82 | 0.998 | theta | T7 | Group | 4.207 | 0.016 | 0.062 | theta |
| T8 | Age | 0.101 | 0.745 | 0.998 | theta | T8 | Group | 0.86 | 0.425 | 0.449 | theta |
| C3 | edu | 0.035 | 0.852 | 0.995 | theta | C3 | sex | 0.504 | 0.471 | 0.901 | theta |
| C4 | edu | 0 | 0.995 | 0.995 | theta | C4 | sex | 0.398 | 0.522 | 0.901 | theta |
| Cz | edu | 0.077 | 0.781 | 0.995 | theta | Cz | sex | 0.266 | 0.605 | 0.901 | theta |
| F3 | edu | 0.282 | 0.596 | 0.995 | theta | F3 | sex | 0.969 | 0.33 | 0.901 | theta |
| F4 | edu | 0.092 | 0.759 | 0.995 | theta | F4 | sex | 0.426 | 0.519 | 0.901 | theta |
| F7 | edu | 0.02 | 0.9 | 0.995 | theta | F7 | sex | 0.329 | 0.569 | 0.901 | theta |
| F8 | edu | 0.003 | 0.954 | 0.995 | theta | F8 | sex | 0.722 | 0.394 | 0.901 | theta |
| Fp1 | edu | 0 | 0.987 | 0.995 | theta | Fp1 | sex | 0.741 | 0.394 | 0.901 | theta |
| Fp2 | edu | 0.003 | 0.956 | 0.995 | theta | Fp2 | sex | 0.843 | 0.353 | 0.901 | theta |
| Fz | edu | 0.065 | 0.792 | 0.995 | theta | Fz | sex | 0.339 | 0.556 | 0.901 | theta |
| O1 | edu | 0.6 | 0.428 | 0.995 | theta | O1 | sex | 0 | 0.991 | 0.991 | theta |
| O2 | edu | 0.001 | 0.98 | 0.995 | theta | O2 | sex | 0.122 | 0.719 | 0.911 | theta |
| P3 | edu | 0.003 | 0.959 | 0.995 | theta | P3 | sex | 0.25 | 0.618 | 0.901 | theta |
| P4 | edu | 0.013 | 0.913 | 0.995 | theta | P4 | sex | 0.047 | 0.824 | 0.92 | theta |
| P7 | edu | 1.714 | 0.191 | 0.995 | theta | P7 | sex | 0.072 | 0.789 | 0.92 | theta |
| P8 | edu | 0.052 | 0.812 | 0.995 | theta | P8 | sex | 0.571 | 0.453 | 0.901 | theta |
| Pz | edu | 0.047 | 0.827 | 0.995 | theta | Pz | sex | 0.001 | 0.978 | 0.991 | theta |
| T7 | edu | 0.927 | 0.338 | 0.995 | theta | T7 | sex | 0.187 | 0.664 | 0.901 | theta |
| T8 | edu | 0.344 | 0.563 | 0.995 | theta | T8 | sex | 2.051 | 0.156 | 0.901 | theta |

**Table S2.** Statistical values of the SLSW theta density, permutation p-values from the ANCOVA, with density as the dependent variable, and Group as the independent variable. Age, sex, and education were used as covariates of no interest.

*Post-hoc comparisons for theta SLSW density*

| chan | effect | F | p |
|---|---|---|---|
| C3 | PD-H vs. PD-nH | 2.455 | 0.122 |
| F3 | PD-H vs. PD-nH | 2.207 | 0.139 |
| Fz | PD-H vs. PD-nH | 1.581 | 0.21 |
| C3 | PD-H vs. HC | 10.93 | 0.002 |
| F3 | PD-H vs. HC | 12.87 | 0.001 |
| Fz | PD-H vs. HC | 10.129 | 0.002 |
| C3 | PD-nH vs. HC | 9.351 | 0.003 |
| F3 | PD-nH vs. HC | 7.86 | 0.006 |
| Fz | PD-nH vs. HC | 6.44 | 0.013 |

**Table S3.** Post-hoc comparisons for the SLSW theta density on the cluster of electrodes showing a significant main effect of Group in Table S2. Post-hoc comparisons models assessed the modulation of density as a function of Group. Age, sex, and education were used as covariates of no interest. Here, p-values are reported for the effect of Group.

*Delta SLSW density is not modulated by PD*

| chan | effect | F | p | pfdr | Band | chan | effect | F | p | pfdr | Band |
|---|---|---|---|---|---|---|---|---|---|---|---|
| C3 | Age | 1.692 | 0.201 | 0.828 | delta | C3 | Group | 0.636 | 0.533 | 0.845 | delta |
| C4 | Age | 1.072 | 0.306 | 0.828 | delta | C4 | Group | 0.036 | 0.964 | 0.964 | delta |
| Cz | Age | 2.602 | 0.112 | 0.828 | delta | Cz | Group | 0.273 | 0.762 | 0.872 | delta |
| F3 | Age | 0.836 | 0.362 | 0.828 | delta | F3 | Group | 0.414 | 0.666 | 0.872 | delta |
| F4 | Age | 1.617 | 0.205 | 0.828 | delta | F4 | Group | 0.931 | 0.398 | 0.769 | delta |
| F7 | Age | 0.053 | 0.826 | 0.828 | delta | F7 | Group | 0.949 | 0.392 | 0.769 | delta |
| F8 | Age | 0.14 | 0.715 | 0.828 | delta | F8 | Group | 1.108 | 0.323 | 0.769 | delta |
| Fp1 | Age | 0.547 | 0.468 | 0.828 | delta | Fp1 | Group | 1.049 | 0.351 | 0.769 | delta |
| Fp2 | Age | 0.171 | 0.685 | 0.828 | delta | Fp2 | Group | 2.366 | 0.096 | 0.769 | delta |
| Fz | Age | 3.307 | 0.073 | 0.828 | delta | Fz | Group | 0.849 | 0.418 | 0.769 | delta |
| O1 | Age | 0.11 | 0.746 | 0.828 | delta | O1 | Group | 1.015 | 0.366 | 0.769 | delta |
| O2 | Age | 0.049 | 0.828 | 0.828 | delta | O2 | Group | 1.237 | 0.301 | 0.769 | delta |
| P3 | Age | 0.522 | 0.466 | 0.828 | delta | P3 | Group | 0.131 | 0.883 | 0.932 | delta |
| P4 | Age | 0.059 | 0.81 | 0.828 | delta | P4 | Group | 0.34 | 0.71 | 0.872 | delta |
| P7 | Age | 0.06 | 0.804 | 0.828 | delta | P7 | Group | 0.846 | 0.442 | 0.769 | delta |
| P8 | Age | 0.137 | 0.711 | 0.828 | delta | P8 | Group | 1.365 | 0.266 | 0.769 | delta |
| Pz | Age | 0.139 | 0.727 | 0.828 | delta | Pz | Group | 0.256 | 0.78 | 0.872 | delta |
| T7 | Age | 0.283 | 0.607 | 0.828 | delta | T7 | Group | 0.436 | 0.647 | 0.872 | delta |
| T8 | Age | 1.102 | 0.286 | 0.828 | delta | T8 | Group | 0.828 | 0.445 | 0.769 | delta |
| C3 | edu | 0.988 | 0.325 | 0.708 | delta | C3 | sex | 0.66 | 0.423 | 0.536 | delta |
| C4 | edu | 0.785 | 0.369 | 0.708 | delta | C4 | sex | 1.553 | 0.223 | 0.536 | delta |
| Cz | edu | 1.264 | 0.263 | 0.708 | delta | Cz | sex | 1.07 | 0.308 | 0.536 | delta |
| F3 | edu | 0.515 | 0.471 | 0.814 | delta | F3 | sex | 0.504 | 0.479 | 0.551 | delta |
| F4 | edu | 0.92 | 0.339 | 0.708 | delta | F4 | sex | 2.389 | 0.122 | 0.536 | delta |
| F7 | edu | 1.028 | 0.317 | 0.708 | delta | F7 | sex | 0.414 | 0.522 | 0.551 | delta |
| F8 | edu | 1.36 | 0.24 | 0.708 | delta | F8 | sex | 0.181 | 0.672 | 0.672 | delta |
| Fp1 | edu | 0.221 | 0.639 | 0.846 | delta | Fp1 | sex | 1.187 | 0.281 | 0.536 | delta |
| Fp2 | edu | 0.26 | 0.624 | 0.846 | delta | Fp2 | sex | 0.73 | 0.395 | 0.536 | delta |
| Fz | edu | 0.417 | 0.528 | 0.837 | delta | Fz | sex | 2.346 | 0.129 | 0.536 | delta |
| O1 | edu | 0.045 | 0.843 | 0.942 | delta | O1 | sex | 1.129 | 0.299 | 0.536 | delta |
| O2 | edu | 0.108 | 0.743 | 0.883 | delta | O2 | sex | 1.567 | 0.218 | 0.536 | delta |
| P3 | edu | 0.004 | 0.945 | 0.945 | delta | P3 | sex | 1.195 | 0.288 | 0.536 | delta |
| P4 | edu | 0.194 | 0.668 | 0.846 | delta | P4 | sex | 0.713 | 0.403 | 0.536 | delta |
| P7 | edu | 0.796 | 0.373 | 0.708 | delta | P7 | sex | 1.027 | 0.312 | 0.536 | delta |
| P8 | edu | 1.637 | 0.212 | 0.708 | delta | P8 | sex | 1.937 | 0.172 | 0.536 | delta |
| Pz | edu | 0.015 | 0.9 | 0.945 | delta | Pz | sex | 0.935 | 0.328 | 0.536 | delta |
| T7 | edu | 0.775 | 0.37 | 0.708 | delta | T7 | sex | 0.42 | 0.516 | 0.551 | delta |
| T8 | edu | 1.28 | 0.262 | 0.708 | delta | T8 | sex | 0.758 | 0.387 | 0.536 | delta |

**Table S4.** Statistical values of the SLSW delta density, permutation p-values from the ANCOVA, with density as the dependent variable, and Group as the independent variable. Age, sex, and education were used as covariates of no interest.

*Fano Factor indicates that the occurrence of SLSW follows a Poisson distribution and SLSW do not occur in clusters*

| Frequency | Group | chan | mean | SD | Group | mean | SD | Group | mean | SD |
|---|---|---|---|---|---|---|---|---|---|---|
| Theta | PD-H | C3 | 0.9946 | 0.0015 | PD-nH | 0.994 | 0.001 | HC | 0.994 | 0.002 |
| Theta | PD-H | C4 | 0.9944 | 0.0014 | PD-nH | 0.994 | 0.001 | HC | 0.994 | 0.002 |
| Theta | PD-H | Cz | 0.9944 | 0.0017 | PD-nH | 0.994 | 0.001 | HC | 0.994 | 0.002 |
| Theta | PD-H | F3 | 0.9943 | 0.0015 | PD-nH | 0.994 | 0.001 | HC | 0.994 | 0.002 |
| Theta | PD-H | F4 | 0.9944 | 0.0015 | PD-nH | 0.994 | 0.001 | HC | 0.994 | 0.002 |
| Theta | PD-H | F7 | 0.9947 | 0.0013 | PD-nH | 0.994 | 0.001 | HC | 0.995 | 0.002 |
| Theta | PD-H | F8 | 0.9946 | 0.0012 | PD-nH | 0.994 | 0.001 | HC | 0.995 | 0.002 |
| Theta | PD-H | Fp1 | 0.9948 | 0.0012 | PD-nH | 0.994 | 0.001 | HC | 0.994 | 0.002 |
| Theta | PD-H | Fp2 | 0.9948 | 0.0013 | PD-nH | 0.994 | 0.001 | HC | 0.994 | 0.002 |
| Theta | PD-H | Fz | 0.9943 | 0.0015 | PD-nH | 0.994 | 0.001 | HC | 0.994 | 0.002 |
| Theta | PD-H | O1 | 0.9946 | 0.0018 | PD-nH | 0.995 | 0.002 | HC | 0.995 | 0.002 |
| Theta | PD-H | O2 | 0.9947 | 0.0017 | PD-nH | 0.995 | 0.002 | HC | 0.995 | 0.002 |
| Theta | PD-H | P3 | 0.9947 | 0.0015 | PD-nH | 0.994 | 0.001 | HC | 0.995 | 0.002 |
| Theta | PD-H | P4 | 0.9946 | 0.0016 | PD-nH | 0.994 | 0.001 | HC | 0.995 | 0.002 |
| Theta | PD-H | P7 | 0.9948 | 0.0015 | PD-nH | 0.995 | 0.001 | HC | 0.995 | 0.002 |
| Theta | PD-H | P8 | 0.9947 | 0.0014 | PD-nH | 0.995 | 0.002 | HC | 0.995 | 0.002 |
| Theta | PD-H | Pz | 0.9946 | 0.0016 | PD-nH | 0.994 | 0.002 | HC | 0.995 | 0.002 |
| Theta | PD-H | T7 | 0.9946 | 0.0013 | PD-nH | 0.994 | 0.001 | HC | 0.995 | 0.001 |
| Theta | PD-H | T8 | 0.9946 | 0.0013 | PD-nH | 0.994 | 0.001 | HC | 0.995 | 0.002 |

| Frequency | Group | chan | mean | SD | Group | mean | SD | Group | mean | SD |
|---|---|---|---|---|---|---|---|---|---|---|
| Delta | PD-H | C3 | 0.995 | 0.001 | PD-nH | 0.994 | 0.001 | HC | 0.994 | 0.002 |
| Delta | PD-H | C4 | 0.994 | 0.001 | PD-nH | 0.994 | 0.001 | HC | 0.994 | 0.002 |
| Delta | PD-H | Cz | 0.994 | 0.002 | PD-nH | 0.994 | 0.001 | HC | 0.994 | 0.002 |
| Delta | PD-H | F3 | 0.994 | 0.002 | PD-nH | 0.994 | 0.001 | HC | 0.994 | 0.002 |
| Delta | PD-H | F4 | 0.994 | 0.002 | PD-nH | 0.994 | 0.001 | HC | 0.994 | 0.002 |
| Delta | PD-H | F7 | 0.995 | 0.001 | PD-nH | 0.994 | 0.001 | HC | 0.995 | 0.002 |
| Delta | PD-H | F8 | 0.995 | 0.001 | PD-nH | 0.994 | 0.001 | HC | 0.995 | 0.002 |
| Delta | PD-H | Fp1 | 0.995 | 0.001 | PD-nH | 0.994 | 0.001 | HC | 0.994 | 0.002 |
| Delta | PD-H | Fp2 | 0.995 | 0.001 | PD-nH | 0.994 | 0.001 | HC | 0.994 | 0.002 |
| Delta | PD-H | Fz | 0.994 | 0.002 | PD-nH | 0.994 | 0.001 | HC | 0.994 | 0.002 |
| Delta | PD-H | O1 | 0.995 | 0.002 | PD-nH | 0.995 | 0.002 | HC | 0.995 | 0.002 |
| Delta | PD-H | O2 | 0.995 | 0.002 | PD-nH | 0.995 | 0.002 | HC | 0.995 | 0.002 |
| Delta | PD-H | P3 | 0.995 | 0.001 | PD-nH | 0.994 | 0.001 | HC | 0.995 | 0.002 |
| Delta | PD-H | P4 | 0.995 | 0.002 | PD-nH | 0.994 | 0.001 | HC | 0.995 | 0.002 |
| Delta | PD-H | P7 | 0.995 | 0.001 | PD-nH | 0.995 | 0.001 | HC | 0.995 | 0.002 |
| Delta | PD-H | P8 | 0.995 | 0.001 | PD-nH | 0.995 | 0.002 | HC | 0.995 | 0.002 |
| Delta | PD-H | Pz | 0.995 | 0.002 | PD-nH | 0.994 | 0.002 | HC | 0.995 | 0.002 |
| Delta | PD-H | T7 | 0.995 | 0.001 | PD-nH | 0.994 | 0.001 | HC | 0.995 | 0.001 |
| Delta | PD-H | T8 | 0.995 | 0.001 | PD-nH | 0.994 | 0.001 | HC | 0.995 | 0.002 |

**Table S5.** The top table indicates the mean Fano Factor (FF) for the theta and delta frequency bands, for each group and channel, as well as the standard deviation (SD).

*Frontal theta SLSW accumulation rate is faster in PD*

We also investigated whether the accumulation rate (i.e., how quickly SLSW events add up over time) (Figure S1A), by comparing among sub-groups the slopes of a fitted linear regression to the cumulative sum of SLSW (computed for each subject and electrode). Our data show a significant (permutation p-values < 0.05, FDR-corrected) difference in accumulation rate between the three groups for the theta SLSW. This difference in accumulation rate between the three subgroups was again localized over fronto-central electrodes (Figure S1B). Post-hoc analyses on significant fronto-central electrodes revealed that PD-H have a more rapid accumulation rate (steeper slope) of SLSW than HC (permutation p-values < 0.05, FDR-corrected; Figure S1B). Furthermore, PD-nH also have a more rapid accumulation rate of theta SLSW than HC (permutation p-values < 0.05, FDR-corrected), and PD-H do not significantly (permutation p-values > 0.05) differ from PD-nH (Figure S1B). There were no significant (all permutation p-values > 0.05, FDR-corrected) differences between the three sub-groups for the delta SLSW (Figure S1C). These results show that theta (but not delta) SLSW accumulate more rapidly in PD patients, compared to HC, linking SLSW accumulation to the neurodegenerative disease.

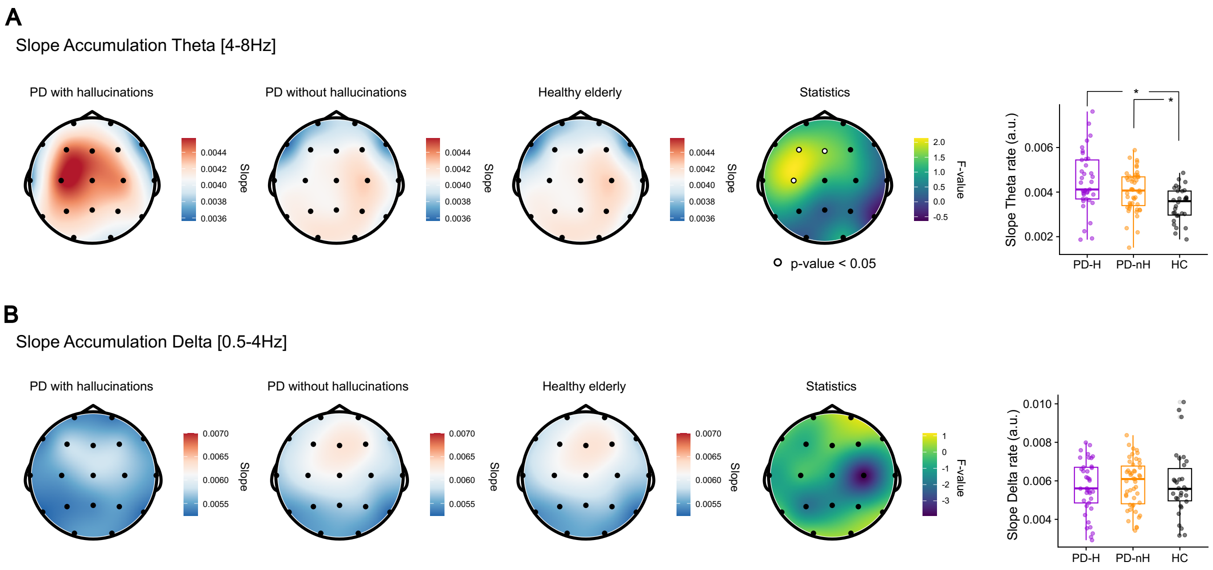


**Figure S1. Theta SLSW accumulate faster in PD with hallucinations. A.** EEG topographies indicating the average across subjects of the slopes of the accumulation rate of theta SLSW obtained for PD-H (left), PD-nH (center) and HC (right). The rightmost topography indicates the F-values of the one-way interaction between the three groups (white dots indicate significant electrodes with permutation p-values < 0.05, FDR-corrected). The box plots show the average density within the significant cluster and the post-hoc results. Single dots represent a single individual. **B.**EEG topographies indicating the average delta SLSW slopes of the accumulation rate obtained for PD-H (left), PD-nH (center), and HC (right). The rightmost topography indicates the F-values of the one-way interaction between the three groups; no significant electrode was observed after FDR correction. The box plot shows the density for the electrode C4 (selected for visualization purposes only). For both sub-plots, each dot represents the average of a single participant.

*Theta SLSW have a higher peak-to-peak amplitude in PD-H*

| chan | effect | F | p | pfdr | Band | chan | effect | F | p | pfdr | Band |
|---|---|---|---|---|---|---|---|---|---|---|---|
| C3 | Age | 8.634 | 0.004 | 0.032 | theta | C3 | Group | 6.030 | 0.003 | 0.007 | theta |
| C4 | Age | 6.856 | 0.010 | 0.032 | theta | C4 | Group | 7.076 | 0.001 | 0.005 | theta |
| Cz | Age | 7.843 | 0.006 | 0.032 | theta | Cz | Group | 7.509 | 0.001 | 0.005 | theta |
| F3 | Age | 7.559 | 0.007 | 0.032 | theta | F3 | Group | 7.744 | 0.001 | 0.005 | theta |
| F4 | Age | 3.718 | 0.056 | 0.083 | theta | F4 | Group | 6.912 | 0.001 | 0.005 | theta |
| F7 | Age | 5.368 | 0.022 | 0.041 | theta | F7 | Group | 5.157 | 0.007 | 0.013 | theta |
| F8 | Age | 0.552 | 0.459 | 0.459 | theta | F8 | Group | 6.244 | 0.003 | 0.006 | theta |
| Fp1 | Age | 1.837 | 0.178 | 0.226 | theta | Fp1 | Group | 6.880 | 0.002 | 0.005 | theta |
| Fp2 | Age | 0.805 | 0.372 | 0.415 | theta | Fp2 | Group | 6.262 | 0.003 | 0.006 | theta |
| Fz | Age | 5.189 | 0.025 | 0.041 | theta | Fz | Group | 7.659 | 0.001 | 0.005 | theta |
| O1 | Age | 5.092 | 0.026 | 0.041 | theta | O1 | Group | 2.799 | 0.065 | 0.073 | theta |
| O2 | Age | 5.958 | 0.016 | 0.041 | theta | O2 | Group | 1.737 | 0.181 | 0.181 | theta |
| P3 | Age | 5.366 | 0.022 | 0.041 | theta | P3 | Group | 4.196 | 0.018 | 0.026 | theta |
| P4 | Age | 5.388 | 0.022 | 0.041 | theta | P4 | Group | 3.135 | 0.047 | 0.056 | theta |
| P7 | Age | 1.449 | 0.231 | 0.275 | theta | P7 | Group | 3.794 | 0.026 | 0.032 | theta |
| P8 | Age | 2.868 | 0.093 | 0.127 | theta | P8 | Group | 1.866 | 0.160 | 0.169 | theta |
| Pz | Age | 7.226 | 0.008 | 0.032 | theta | Pz | Group | 4.396 | 0.015 | 0.023 | theta |
| T7 | Age | 7.348 | 0.008 | 0.032 | theta | T7 | Group | 5.793 | 0.004 | 0.008 | theta |
| T8 | Age | 0.644 | 0.424 | 0.448 | theta | T8 | Group | 4.015 | 0.021 | 0.028 | theta |
| C3 | edu | 0.132 | 0.717 | 0.962 | theta | C3 | sex | 11.593 | 0.001 | 0.003 | theta |
| C4 | edu | 0.040 | 0.843 | 0.962 | theta | C4 | sex | 8.171 | 0.005 | 0.010 | theta |
| Cz | edu | 0.179 | 0.673 | 0.962 | theta | Cz | sex | 6.967 | 0.010 | 0.016 | theta |
| F3 | edu | 0.532 | 0.467 | 0.962 | theta | F3 | sex | 5.360 | 0.022 | 0.032 | theta |
| F4 | edu | 0.474 | 0.492 | 0.962 | theta | F4 | sex | 5.290 | 0.023 | 0.032 | theta |
| F7 | edu | 1.000 | 0.320 | 0.962 | theta | F7 | sex | 3.658 | 0.058 | 0.069 | theta |
| F8 | edu | 0.157 | 0.692 | 0.962 | theta | F8 | sex | 1.502 | 0.223 | 0.223 | theta |
| Fp1 | edu | 0.338 | 0.562 | 0.962 | theta | Fp1 | sex | 1.685 | 0.197 | 0.208 | theta |
| Fp2 | edu | 0.245 | 0.621 | 0.962 | theta | Fp2 | sex | 2.486 | 0.118 | 0.132 | theta |
| Fz | edu | 0.479 | 0.491 | 0.962 | theta | Fz | sex | 5.057 | 0.027 | 0.034 | theta |
| O1 | edu | 0.040 | 0.842 | 0.962 | theta | O1 | sex | 15.865 | 0.000 | 0.001 | theta |
| O2 | edu | 0.049 | 0.826 | 0.962 | theta | O2 | sex | 19.484 | 0.000 | 0.000 | theta |
| P3 | edu | 0.002 | 0.962 | 0.962 | theta | P3 | sex | 15.313 | 0.000 | 0.001 | theta |
| P4 | edu | 0.015 | 0.902 | 0.962 | theta | P4 | sex | 13.899 | 0.000 | 0.001 | theta |
| P7 | edu | 0.505 | 0.479 | 0.962 | theta | P7 | sex | 14.216 | 0.000 | 0.001 | theta |
| P8 | edu | 0.059 | 0.809 | 0.962 | theta | P8 | sex | 11.048 | 0.001 | 0.003 | theta |
| Pz | edu | 0.013 | 0.908 | 0.962 | theta | Pz | sex | 11.078 | 0.001 | 0.003 | theta |
| T7 | edu | 0.438 | 0.509 | 0.962 | theta | T7 | sex | 9.031 | 0.003 | 0.007 | theta |
| T8 | edu | 0.006 | 0.939 | 0.962 | theta | T8 | sex | 5.261 | 0.024 | 0.032 | theta |

**Table S6.** Statistical values of the SLSW theta peak-to-peak amplitude difference between groups, p-values from the linear mixed models, with amplitude as the dependent variable, and Group as the fixed effect. Age, sex, and education were used as covariates of no interest.

*Post-hoc comparisons for theta SLSW higher peak-to-peak amplitude*

| chan | effect | F | p | chan | effect | F | p | chan | effect | F | p |
|---|---|---|---|---|---|---|---|---|---|---|---|
| C3 | PD-H vs. PD-nH | 6.146 | 0.015 | C3 | PD-H vs. HC | 10.289 | 0.002 | C3 | PD-nH vs. HC | 0.895 | 0.347 |
| C4 | PD-H vs. PD-nH | 7.374 | 0.008 | C4 | PD-H vs. HC | 12.872 | 0.001 | C4 | PD-nH vs. HC | 0.826 | 0.366 |
| Cz | PD-H vs. PD-nH | 6.246 | 0.015 | Cz | PD-H vs. HC | 16.268 | 0.000 | Cz | PD-nH vs. HC | 1.695 | 0.197 |
| F3 | PD-H vs. PD-nH | 7.992 | 0.006 | F3 | PD-H vs. HC | 12.641 | 0.001 | F3 | PD-nH vs. HC | 1.294 | 0.259 |
| F4 | PD-H vs. PD-nH | 6.701 | 0.011 | F4 | PD-H vs. HC | 11.962 | 0.001 | F4 | PD-nH vs. HC | 1.307 | 0.257 |
| F7 | PD-H vs. PD-nH | 4.842 | 0.031 | F7 | PD-H vs. HC | 8.389 | 0.005 | F7 | PD-nH vs. HC | 1.145 | 0.288 |
| F8 | PD-H vs. PD-nH | 6.949 | 0.010 | F8 | PD-H vs. HC | 9.776 | 0.003 | F8 | PD-nH vs. HC | 0.865 | 0.356 |
| Fp1 | PD-H vs. PD-nH | 5.253 | 0.025 | Fp1 | PD-H vs. HC | 13.579 | 0.000 | Fp1 | PD-nH vs. HC | 2.287 | 0.135 |
| Fp2 | PD-H vs. PD-nH | 5.543 | 0.021 | Fp2 | PD-H vs. HC | 11.408 | 0.001 | Fp2 | PD-nH vs. HC | 1.546 | 0.218 |
| Fz | PD-H vs. PD-nH | 6.994 | 0.010 | Fz | PD-H vs. HC | 13.592 | 0.000 | Fz | PD-nH vs. HC | 1.722 | 0.194 |
| P3 | PD-H vs. PD-nH | 4.034 | 0.048 | P3 | PD-H vs. HC | 7.492 | 0.008 | P3 | PD-nH vs. HC | 0.626 | 0.431 |
| P7 | PD-H vs. PD-nH | 5.109 | 0.027 | P7 | PD-H vs. HC | 4.108 | 0.047 | P7 | PD-nH vs. HC | 0.000 | 0.996 |
| Pz | PD-H vs. PD-nH | 3.232 | 0.076 | Pz | PD-H vs. HC | 10.748 | 0.002 | Pz | PD-nH vs. HC | 1.224 | 0.272 |
| T7 | PD-H vs. PD-nH | 7.864 | 0.006 | T7 | PD-H vs. HC | 5.825 | 0.019 | T7 | PD-nH vs. HC | 0.109 | 0.742 |
| T8 | PD-H vs. PD-nH | 5.790 | 0.018 | T8 | PD-H vs. HC | 4.315 | 0.042 | T8 | PD-nH vs. HC | 0.141 | 0.708 |

**Table S7.** Theta post-hoc comparisons for the SLSW theta peak-to-peak on the electrodes showing a significant main effect of Group in Table S6. Post-hoc comparisons models assessed the modulation of density as a function of Group (permutation p-values). Age, sex, and education were used as covariates of no interest. Here, p-values are reported for the effect of Group.

*Delta SLSW have higher peak-to-peak amplitude in PD-H*

| chan | effect | F | p | pfdr | chan | effect | F | p | pfdr |
|---|---|---|---|---|---|---|---|---|---|
| C3 | Age | 8.634 | 0.004 | 0.032 | C3 | Group | 6.029519 | 0.003289 | 0.006943 |
| C4 | Age | 6.856 | 0.010 | 0.032 | C4 | Group | 7.076 | 0.001 | 0.005 |
| Cz | Age | 7.843 | 0.006 | 0.032 | Cz | Group | 7.509 | 0.001 | 0.005 |
| F3 | Age | 7.559 | 0.007 | 0.032 | F3 | Group | 7.744 | 0.001 | 0.005 |
| F4 | Age | 3.718 | 0.056 | 0.083 | F4 | Group | 6.912 | 0.001 | 0.005 |
| F7 | Age | 5.368 | 0.022 | 0.041 | F7 | Group | 5.157 | 0.007 | 0.013 |
| F8 | Age | 0.552 | 0.459 | 0.459 | F8 | Group | 6.244 | 0.003 | 0.006 |
| Fp1 | Age | 1.837 | 0.178 | 0.226 | Fp1 | Group | 6.880 | 0.002 | 0.005 |
| Fp2 | Age | 0.805 | 0.372 | 0.415 | Fp2 | Group | 6.262 | 0.003 | 0.006 |
| Fz | Age | 5.189 | 0.025 | 0.041 | Fz | Group | 7.659 | 0.001 | 0.005 |
| O1 | Age | 5.092 | 0.026 | 0.041 | O1 | Group | 2.799 | 0.065 | 0.073 |
| O2 | Age | 5.958 | 0.016 | 0.041 | O2 | Group | 1.737 | 0.181 | 0.181 |
| P3 | Age | 5.366 | 0.022 | 0.041 | P3 | Group | 4.196 | 0.018 | 0.026 |
| P4 | Age | 5.388 | 0.022 | 0.041 | P4 | Group | 3.135 | 0.047 | 0.056 |
| P7 | Age | 1.449 | 0.231 | 0.275 | P7 | Group | 3.794 | 0.026 | 0.032 |
| P8 | Age | 2.868 | 0.093 | 0.127 | P8 | Group | 1.866 | 0.160 | 0.169 |
| Pz | Age | 7.226 | 0.008 | 0.032 | Pz | Group | 4.396 | 0.015 | 0.023 |
| T7 | Age | 7.348 | 0.008 | 0.032 | T7 | Group | 5.793 | 0.004 | 0.008 |
| T8 | Age | 0.644 | 0.424 | 0.448 | T8 | Group | 4.015 | 0.021 | 0.028 |
| C3 | edu | 0.132 | 0.717 | 0.962 | C3 | sex | 11.593 | 0.001 | 0.003 |
| C4 | edu | 0.040 | 0.843 | 0.962 | C4 | sex | 8.171 | 0.005 | 0.010 |
| Cz | edu | 0.179 | 0.673 | 0.962 | Cz | sex | 6.967 | 0.010 | 0.016 |
| F3 | edu | 0.532 | 0.467 | 0.962 | F3 | sex | 5.360 | 0.022 | 0.032 |
| F4 | edu | 0.474 | 0.492 | 0.962 | F4 | sex | 5.290 | 0.023 | 0.032 |
| F7 | edu | 1.000 | 0.320 | 0.962 | F7 | sex | 3.658 | 0.058 | 0.069 |
| F8 | edu | 0.157 | 0.692 | 0.962 | F8 | sex | 1.502 | 0.223 | 0.223 |
| Fp1 | edu | 0.338 | 0.562 | 0.962 | Fp1 | sex | 1.685 | 0.197 | 0.208 |
| Fp2 | edu | 0.245 | 0.621 | 0.962 | Fp2 | sex | 2.486 | 0.118 | 0.132 |
| Fz | edu | 0.479 | 0.491 | 0.962 | Fz | sex | 5.057 | 0.027 | 0.034 |
| O1 | edu | 0.040 | 0.842 | 0.962 | O1 | sex | 15.865 | 0.000 | 0.001 |
| O2 | edu | 0.049 | 0.826 | 0.962 | O2 | sex | 19.484 | 0.000 | 0.000 |
| P3 | edu | 0.002 | 0.962 | 0.962 | P3 | sex | 15.313 | 0.000 | 0.001 |
| P4 | edu | 0.015 | 0.902 | 0.962 | P4 | sex | 13.899 | 0.000 | 0.001 |
| P7 | edu | 0.505 | 0.479 | 0.962 | P7 | sex | 14.216 | 0.000 | 0.001 |
| P8 | edu | 0.059 | 0.809 | 0.962 | P8 | sex | 11.048 | 0.001 | 0.003 |
| Pz | edu | 0.013 | 0.908 | 0.962 | Pz | sex | 11.078 | 0.001 | 0.003 |
| T7 | edu | 0.438 | 0.509 | 0.962 | T7 | sex | 9.031 | 0.003 | 0.007 |
| T8 | edu | 0.006 | 0.939 | 0.962 | T8 | sex | 5.261 | 0.024 | 0.032 |

**Table S8.** Statistical values of the SLSW delta peak-to-peak amplitude difference between groups, p-values from the linear mixed models, with amplitude as the dependent variable, and Group as the fixed effect. Age, sex, and education were used as covariates of no interest.

### *Post-hoc comparisons for delta SLSW higher peak-to-peak amplitude*

| chan | effect | F | p | chan | effect | F | p | chan | effect | F | p |
|---|---|---|---|---|---|---|---|---|---|---|---|
| C3 | PD-H vs. PD-nH | 5.448 | 0.022 | C3 | PD-H vs. HC | 8.831 | 0.004 | C3 | PD-nH vs. HC | 0.538 | 0.466 |
| C4 | PD-H vs. PD-nH | 9.412 | 0.003 | C4 | PD-H vs. HC | 12.676 | 0.001 | C4 | PD-nH vs. HC | 0.172 | 0.680 |
| Cz | PD-H vs. PD-nH | 6.636 | 0.012 | Cz | PD-H vs. HC | 15.389 | 0.000 | Cz | PD-nH vs. HC | 0.815 | 0.370 |
| F3 | PD-H vs. PD-nH | 5.671 | 0.020 | F3 | PD-H vs. HC | 7.886 | 0.007 | F3 | PD-nH vs. HC | 0.683 | 0.411 |
| F4 | PD-H vs. PD-nH | 6.694 | 0.012 | F4 | PD-H vs. HC | 11.066 | 0.001 | F4 | PD-nH vs. HC | 0.668 | 0.416 |
| F7 | PD-H vs. PD-nH | 6.873 | 0.010 | F7 | PD-H vs. HC | 7.779 | 0.007 | F7 | PD-nH vs. HC | 0.407 | 0.525 |
| F8 | PD-H vs. PD-nH | 9.064 | 0.003 | F8 | PD-H vs. HC | 7.285 | 0.009 | F8 | PD-nH vs. HC | 0.206 | 0.652 |
| Fp1 | PD-H vs. PD-nH | 5.980 | 0.017 | Fp1 | PD-H vs. HC | 8.842 | 0.004 | Fp1 | PD-nH vs. HC | 1.270 | 0.264 |
| Fp2 | PD-H vs. PD-nH | 7.284 | 0.009 | Fp2 | PD-H vs. HC | 9.049 | 0.004 | Fp2 | PD-nH vs. HC | 0.647 | 0.424 |
| Fz | PD-H vs. PD-nH | 7.074 | 0.009 | Fz | PD-H vs. HC | 12.926 | 0.001 | Fz | PD-nH vs. HC | 1.190 | 0.279 |
| O1 | PD-H vs. PD-nH | 7.701 | 0.007 | O1 | PD-H vs. HC | 4.733 | 0.033 | O1 | PD-nH vs. HC | 0.362 | 0.549 |
| O2 | PD-H vs. PD-nH | 5.240 | 0.025 | O2 | PD-H vs. HC | 3.242 | 0.077 | O2 | PD-nH vs. HC | 0.193 | 0.662 |
| P3 | PD-H vs. PD-nH | 4.293 | 0.042 | P3 | PD-H vs. HC | 6.498 | 0.013 | P3 | PD-nH vs. HC | 0.447 | 0.506 |
| P4 | PD-H vs. PD-nH | 5.197 | 0.025 | P4 | PD-H vs. HC | 8.393 | 0.005 | P4 | PD-nH vs. HC | 0.391 | 0.534 |
| P7 | PD-H vs. PD-nH | 5.448 | 0.022 | P7 | PD-H vs. HC | 3.248 | 0.076 | P7 | PD-nH vs. HC | 0.036 | 0.851 |
| Pz | PD-H vs. PD-nH | 4.853 | 0.031 | Pz | PD-H vs. HC | 11.611 | 0.001 | Pz | PD-nH vs. HC | 0.941 | 0.335 |
| T7 | PD-H vs. PD-nH | 7.186 | 0.009 | T7 | PD-H vs. HC | 2.927 | 0.092 | T7 | PD-nH vs. HC | 0.072 | 0.789 |
| T8 | PD-H vs. PD-nH | 8.226 | 0.005 | T8 | PD-H vs. HC | 4.510 | 0.038 | T8 | PD-nH vs. HC | 0.011 | 0.918 |

**Table S9.** Delta peak-to-peak post-hoc comparisons on the significant clusters shown in Figure 2. Here, in the linear mixed models, we also control for disease duration and daily levodopa equivalent intake, in addition to age, education, and sex.

*SLSW downward slope is steeper in PD-H compared to PD-nH and HC*

We analysed whether the downward slopes were modulated differently between the three subgroups (PD-H, PD-nH, and HC). Our results show that for both delta and theta slow waves, there is significant interaction (p-values < 0.05; FDR-corrected), over fronto-central regions (Figure S2, Table 3).

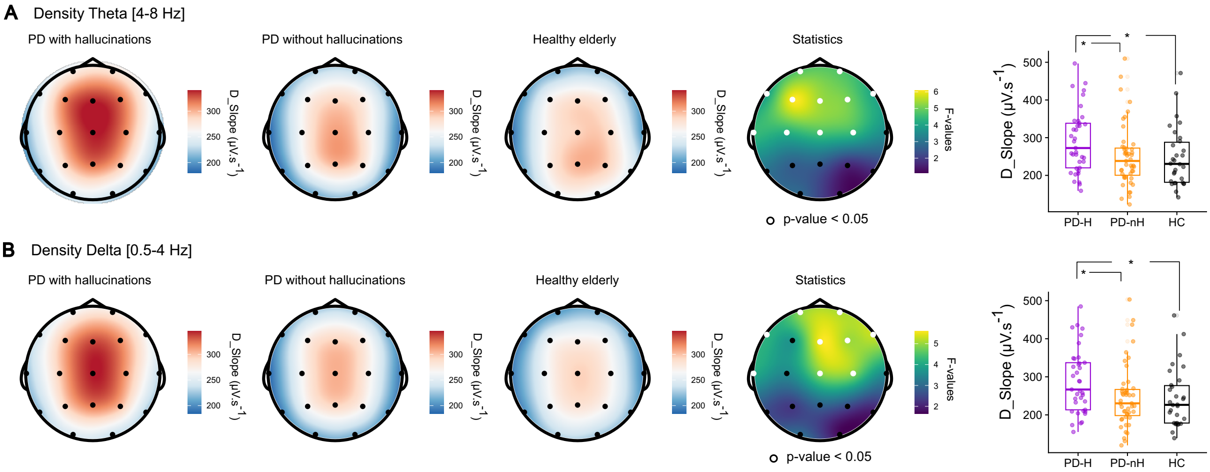


**Figure S2. SLSW downward slope is higher in PD-H than in PD-nH and HC. A**. Theta SLSW downward slope topographies, with topographies indicating the average across subjects. The rightmost topography indicates the F-values of the one-way interaction between the three groups (white dots indicate p-values < 0.05, FDR-corrected). The box plots indicate post-hoc results analyses of the significant fronto-central cluster; each dot represents the average of a single participant. **B**. Delta SLSW downward slope topographies, with topographies indicating the average across subjects. The rightmost topography indicates the F-values of the one-way interaction between the three groups (white dots indicate p-values < 0.05, FDR-corrected). The box plots indicate post-hoc results analyses of the significant fronto-central cluster; each dot represents the average of a single participant.

*SLSW theta downward slope*

| chan | effect | F | p | pfdr | chan | effect | F | p | pfdr |
|---|---|---|---|---|---|---|---|---|---|
| C3 | Age | 6.106 | 0.015 | 0.059 | C3 | Group | 4.256 | 0.017 | 0.035 |
| C4 | Age | 5.155 | 0.025 | 0.061 | C4 | Group | 4.489 | 0.013 | 0.034 |
| Cz | Age | 4.797 | 0.031 | 0.061 | Cz | Group | 4.668 | 0.011 | 0.034 |
| F3 | Age | 7.440 | 0.007 | 0.059 | F3 | Group | 6.058 | 0.003 | 0.034 |
| F4 | Age | 4.258 | 0.041 | 0.061 | F4 | Group | 5.059 | 0.008 | 0.034 |
| F7 | Age | 4.633 | 0.034 | 0.061 | F7 | Group | 3.936 | 0.022 | 0.039 |
| F8 | Age | 0.331 | 0.566 | 0.566 | F8 | Group | 4.065 | 0.020 | 0.038 |
| Fp1 | Age | 1.744 | 0.189 | 0.225 | Fp1 | Group | 4.790 | 0.010 | 0.034 |
| Fp2 | Age | 0.839 | 0.362 | 0.404 | Fp2 | Group | 4.738 | 0.011 | 0.034 |
| Fz | Age | 4.677 | 0.033 | 0.061 | Fz | Group | 5.358 | 0.006 | 0.034 |
| O1 | Age | 4.383 | 0.039 | 0.061 | O1 | Group | 2.786 | 0.066 | 0.090 |
| O2 | Age | 5.798 | 0.018 | 0.059 | O2 | Group | 1.375 | 0.257 | 0.257 |
| P3 | Age | 6.389 | 0.013 | 0.059 | P3 | Group | 2.614 | 0.078 | 0.095 |
| P4 | Age | 4.448 | 0.037 | 0.061 | P4 | Group | 1.654 | 0.196 | 0.207 |
| P7 | Age | 1.833 | 0.179 | 0.225 | P7 | Group | 2.992 | 0.054 | 0.079 |
| P8 | Age | 2.782 | 0.098 | 0.133 | P8 | Group | 1.732 | 0.182 | 0.203 |
| Pz | Age | 6.933 | 0.010 | 0.059 | Pz | Group | 2.589 | 0.080 | 0.095 |
| T7 | Age | 5.720 | 0.018 | 0.059 | T7 | Group | 4.425 | 0.014 | 0.034 |
| T8 | Age | 0.656 | 0.420 | 0.443 | T8 | Group | 3.526 | 0.033 | 0.052 |
| C3 | edu | 0.249 | 0.619 | 0.944 | C3 | sex | 9.717 | 0.002 | 0.006 |
| C4 | edu | 0.121 | 0.729 | 0.944 | C4 | sex | 8.274 | 0.005 | 0.009 |
| Cz | edu | 0.220 | 0.640 | 0.944 | Cz | sex | 4.936 | 0.028 | 0.042 |
| F3 | edu | 0.520 | 0.472 | 0.944 | F3 | sex | 4.932 | 0.028 | 0.042 |
| F4 | edu | 0.609 | 0.437 | 0.944 | F4 | sex | 4.643 | 0.033 | 0.045 |
| F7 | edu | 1.446 | 0.232 | 0.944 | F7 | sex | 3.179 | 0.077 | 0.092 |
| F8 | edu | 0.272 | 0.603 | 0.944 | F8 | sex | 1.529 | 0.219 | 0.219 |
| Fp1 | edu | 0.336 | 0.563 | 0.944 | Fp1 | sex | 1.920 | 0.169 | 0.178 |
| Fp2 | edu | 0.213 | 0.645 | 0.944 | Fp2 | sex | 2.584 | 0.111 | 0.124 |
| Fz | edu | 0.404 | 0.527 | 0.944 | Fz | sex | 4.227 | 0.042 | 0.053 |
| O1 | edu | 0.020 | 0.888 | 0.944 | O1 | sex | 13.033 | 0.000 | 0.002 |
| O2 | edu | 0.018 | 0.894 | 0.944 | O2 | sex | 17.391 | 0.000 | 0.001 |
| P3 | edu | 0.070 | 0.791 | 0.944 | P3 | sex | 15.688 | 0.000 | 0.001 |
| P4 | edu | 0.047 | 0.829 | 0.944 | P4 | sex | 13.142 | 0.000 | 0.002 |
| P7 | edu | 0.560 | 0.456 | 0.944 | P7 | sex | 14.033 | 0.000 | 0.002 |
| P8 | edu | 0.149 | 0.700 | 0.944 | P8 | sex | 10.788 | 0.001 | 0.004 |
| Pz | edu | 0.064 | 0.801 | 0.944 | Pz | sex | 10.863 | 0.001 | 0.004 |
| T7 | edu | 0.635 | 0.427 | 0.944 | T7 | sex | 9.167 | 0.003 | 0.007 |
| T8 | edu | 0.002 | 0.965 | 0.965 | T8 | sex | 6.657 | 0.011 | 0.019 |

**Table S10.** Statistical values of the SLSW theta downward slope difference (Figure S2A) between groups, with amplitude as the dependent variable and Group as the fixed effect (p-values from the linear mixed models). Age, sex, and education were used as covariates of no interest.

*SLSW theta downward slope post-hoc comparisons*

| chan | effect | F | p | chan | effect | F | p | chan | effect | F | p |
|---|---|---|---|---|---|---|---|---|---|---|---|
| C3 | PD-H vs. PD-nH | 5.806 | 0.018 | C3 | PD-H vs HC | 5.903 | 0.018 | C3 | PD-nH vs. HC | 0.123 | 0.727 |
| C4 | PD-H vs. PD-nH | 6.107 | 0.016 | C4 | PD-H vs HC | 6.238 | 0.015 | C4 | PD-nH vs. HC | 0.036 | 0.851 |
| Cz | PD-H vs. PD-nH | 4.299 | 0.041 | Cz | PD-H vs HC | 9.809 | 0.003 | Cz | PD-nH vs. HC | 0.751 | 0.389 |
| F3 | PD-H vs. PD-nH | 7.374 | 0.008 | F3 | PD-H vs HC | 9.258 | 0.003 | F3 | PD-nH vs. HC | 0.529 | 0.470 |
| F4 | PD-H vs. PD-nH | 6.004 | 0.016 | F4 | PD-H vs HC | 8.227 | 0.006 | F4 | PD-nH vs. HC | 0.343 | 0.560 |
| F7 | PD-H vs. PD-nH | 4.251 | 0.043 | F7 | PD-H vs HC | 6.727 | 0.012 | F7 | PD-nH vs. HC | 0.351 | 0.555 |
| F8 | PD-H vs. PD-nH | 5.700 | 0.019 | F8 | PD-H vs HC | 5.692 | 0.020 | F8 | PD-nH vs. HC | 0.075 | 0.786 |
| Fp1 | PD-H vs. PD-nH | 4.195 | 0.044 | Fp1 | PD-H vs HC | 10.028 | 0.002 | Fp1 | PD-nH vs. HC | 0.961 | 0.330 |
| Fp2 | PD-H vs. PD-nH | 4.705 | 0.033 | Fp2 | PD-H vs HC | 9.265 | 0.003 | Fp2 | PD-nH vs. HC | 0.607 | 0.438 |
| Fz | PD-H vs. PD-nH | 5.657 | 0.020 | Fz | PD-H vs HC | 9.390 | 0.003 | Fz | PD-nH vs. HC | 0.569 | 0.453 |
| T7 | PD-H vs. PD-nH | 6.892 | 0.010 | T7 | PD-H vs HC | 4.056 | 0.048 | T7 | PD-nH vs. HC | 0.001 | 0.980 |

**Table S11.** SLSW downward slope post-hoc comparisons on the significant clusters shown in Figure S2A. Here, in the linear mixed models, we also control for age, education, and sex.

*SLSW delta downward slope*

| chan | effect | F | p | pfdr | chan | effect | F | p | pfdr |
|---|---|---|---|---|---|---|---|---|---|
| C3 | Age | 5.241 | 0.024 | 0.129 | C3 | Group | 3.523 | 0.033 | 0.052 |
| C4 | Age | 3.956 | 0.049 | 0.129 | C4 | Group | 4.852 | 0.010 | 0.026 |
| Cz | Age | 4.642 | 0.033 | 0.129 | Cz | Group | 4.846 | 0.010 | 0.026 |
| F3 | Age | 4.605 | 0.034 | 0.129 | F3 | Group | 3.761 | 0.026 | 0.050 |
| F4 | Age | 2.356 | 0.128 | 0.202 | F4 | Group | 5.035 | 0.008 | 0.026 |
| F7 | Age | 4.358 | 0.039 | 0.129 | F7 | Group | 4.174 | 0.018 | 0.043 |
| F8 | Age | 0.130 | 0.719 | 0.719 | F8 | Group | 5.284 | 0.006 | 0.026 |
| Fp1 | Age | 1.524 | 0.220 | 0.278 | Fp1 | Group | 4.901 | 0.009 | 0.026 |
| Fp2 | Age | 0.396 | 0.530 | 0.593 | Fp2 | Group | 5.108 | 0.008 | 0.026 |
| Fz | Age | 3.377 | 0.069 | 0.129 | Fz | Group | 5.541 | 0.005 | 0.026 |
| O1 | Age | 3.245 | 0.074 | 0.129 | O1 | Group | 3.071 | 0.050 | 0.074 |
| O2 | Age | 5.110 | 0.026 | 0.129 | O2 | Group | 1.754 | 0.178 | 0.178 |
| P3 | Age | 1.802 | 0.182 | 0.254 | P3 | Group | 2.325 | 0.103 | 0.115 |
| P4 | Age | 3.346 | 0.070 | 0.129 | P4 | Group | 2.495 | 0.087 | 0.104 |
| P7 | Age | 1.297 | 0.257 | 0.306 | P7 | Group | 2.569 | 0.081 | 0.103 |
| P8 | Age | 1.760 | 0.187 | 0.254 | P8 | Group | 1.932 | 0.150 | 0.158 |
| Pz | Age | 3.537 | 0.063 | 0.129 | Pz | Group | 2.900 | 0.059 | 0.081 |
| T7 | Age | 4.098 | 0.045 | 0.129 | T7 | Group | 4.022 | 0.021 | 0.044 |
| T8 | Age | 0.252 | 0.617 | 0.651 | T8 | Group | 3.519 | 0.033 | 0.052 |
| C3 | edu | 0.250 | 0.618 | 0.943 | C3 | sex | 10.065 | 0.002 | 0.005 |
| C4 | edu | 0.007 | 0.932 | 0.965 | C4 | sex | 8.456 | 0.004 | 0.008 |
| Cz | edu | 0.024 | 0.876 | 0.965 | Cz | sex | 6.326 | 0.013 | 0.021 |
| F3 | edu | 0.603 | 0.439 | 0.943 | F3 | sex | 3.346 | 0.070 | 0.089 |
| F4 | edu | 0.113 | 0.738 | 0.943 | F4 | sex | 4.582 | 0.035 | 0.051 |
| F7 | edu | 0.740 | 0.391 | 0.943 | F7 | sex | 2.432 | 0.122 | 0.138 |
| F8 | edu | 0.107 | 0.745 | 0.943 | F8 | sex | 1.310 | 0.255 | 0.255 |
| Fp1 | edu | 0.344 | 0.559 | 0.943 | Fp1 | sex | 1.540 | 0.217 | 0.229 |
| Fp2 | edu | 0.148 | 0.701 | 0.943 | Fp2 | sex | 2.404 | 0.124 | 0.138 |
| Fz | edu | 0.337 | 0.563 | 0.943 | Fz | sex | 4.030 | 0.047 | 0.064 |
| O1 | edu | 0.552 | 0.459 | 0.943 | O1 | sex | 15.196 | 0.000 | 0.001 |
| O2 | edu | 0.217 | 0.642 | 0.943 | O2 | sex | 18.568 | 0.000 | 0.001 |
| P3 | edu | 0.198 | 0.657 | 0.943 | P3 | sex | 16.551 | 0.000 | 0.001 |
| P4 | edu | 0.117 | 0.733 | 0.943 | P4 | sex | 13.359 | 0.000 | 0.002 |
| P7 | edu | 0.483 | 0.489 | 0.943 | P7 | sex | 14.657 | 0.000 | 0.001 |
| P8 | edu | 0.220 | 0.640 | 0.943 | P8 | sex | 11.582 | 0.001 | 0.003 |
| Pz | edu | 0.002 | 0.965 | 0.965 | Pz | sex | 10.328 | 0.002 | 0.005 |
| T7 | edu | 0.150 | 0.699 | 0.943 | T7 | sex | 8.709 | 0.004 | 0.008 |
| T8 | edu | 0.024 | 0.877 | 0.965 | T8 | sex | 6.881 | 0.010 | 0.017 |

**Table S12.** Statistical values of the SLSW theta downward slope difference (Figure S2B) between groups (p-values from the linear mixed models), with amplitude as the dependent variable, and Group as the fixed effect. Age, sex, and education were used as covariates of no interest.

### *SLSW delta downward slope post-hoc comparisons*

| chan | effect | F | p | chan | effect | F | p | chan | effect | F | p |
|---|---|---|---|---|---|---|---|---|---|---|---|
| C4 | PD-H vs. PD-nH | 6.841 | 0.011 | C4 | PD-H vs. HC | 6.932 | 0.011 | C4 | PD-nH vs. HC | 8.97E-05 | 0.992 |
| Cz | PD-H vs. PD-nH | 5.094 | 0.027 | Cz | PD-H vs. HC | 10.056 | 0.002 | Cz | PD-nH vs. HC | 0.258 | 0.613 |
| F4 | PD-H vs. PD-nH | 5.483 | 0.022 | F4 | PD-H vs. HC | 8.488 | 0.005 | F4 | PD-nH vs. HC | 0.424 | 0.517 |
| F7 | PD-H vs. PD-nH | 4.585 | 0.035 | F7 | PD-H vs. HC | 6.393 | 0.014 | F7 | PD-nH vs. HC | 0.415 | 0.521 |
| F8 | PD-H vs. PD-nH | 7.826 | 0.006 | F8 | PD-H vs. HC | 5.976 | 0.017 | F8 | PD-nH vs. HC | 0.005 | 0.947 |
| Fp1 | PD-H vs. PD-nH | 4.286 | 0.042 | Fp1 | PD-H vs. HC | 9.447 | 0.003 | Fp1 | PD-nH vs. HC | 1.218 | 0.273 |
| Fp2 | PD-H vs. PD-nH | 5.977 | 0.017 | Fp2 | PD-H vs. HC | 7.988 | 0.006 | Fp2 | PD-nH vs. HC | 0.262 | 0.610 |
| Fz | PD-H vs. PD-nH | 5.257 | 0.025 | Fz | PD-H vs. HC | 10.490 | 0.002 | Fz | PD-nH vs. HC | 0.920 | 0.341 |
| T7 | PD-H vs. PD-nH | 6.609 | 0.012 | T7 | PD-H vs. HC | 2.874 | 0.095 | T7 | PD-nH vs. HC | 0.130 | 0.720 |

**Table S13.** SLSW delta downward slope post-hoc comparisons on the significant clusters shown in Figure S2B. Here, in the linear mixed models, we also control for age, education, and sex.

### *SLSW upward slope is steeper in PD-H compared to PD-nH and HC*

We analysed whether the upward slopes were modulated differently between the three subgroups (PD-H, PD-nH, and HC). Our results show that for both delta and theta slow waves there is significant interaction (p-values < 0.05; FDR-corrected), over fronto-central regions (Figure S3, Table S4).

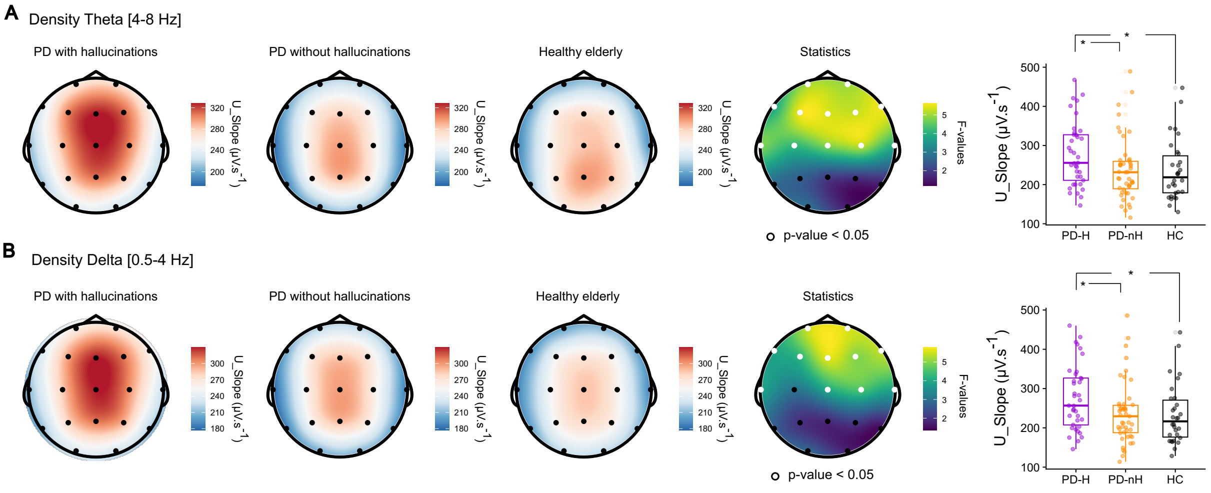


**Figure S3. SLSW upward slope is higher in PD-H than in PD-nH and HC. A**. Theta SLSW upward slope topographies, with topographies indicating the average across subjects. The right topography indicates the F-values of the one-way interaction between the three groups (white dots indicate p-values < 0.05, FDR-corrected). The box plots indicate post-hoc results analyses of the significant fronto-central cluster, each dot represents the average of a single participant. **B**. Delta SLSW upward slope topographies, with topographies indicating the average across subjects. The rightmost topography indicates the F-values of the one-way interaction between the three groups (white dots indicate p-values < 0.05, FDR-corrected). The box plots indicate post-hoc results analyses of the significant fronto-central cluster; each dot represents the average of a single participant.

*SLSW theta upward slope*

| chan | effect | F | p | pfdr | chan | effect | F | p | pfdr |
|---|---|---|---|---|---|---|---|---|---|
| C3 | Age | 7.312 | 0.008 | 0.060 | C3 | Group | 4.148 | 0.018 | 0.035 |
| C4 | Age | 4.909 | 0.029 | 0.066 | C4 | Group | 5.223 | 0.007 | 0.022 |
| Cz | Age | 6.279 | 0.014 | 0.060 | Cz | Group | 4.836 | 0.010 | 0.022 |
| F3 | Age | 6.010 | 0.016 | 0.060 | F3 | Group | 5.526 | 0.005 | 0.022 |
| F4 | Age | 3.319 | 0.071 | 0.101 | F4 | Group | 5.249 | 0.007 | 0.022 |
| F7 | Age | 4.448 | 0.037 | 0.066 | F7 | Group | 3.798 | 0.025 | 0.040 |
| F8 | Age | 0.420 | 0.518 | 0.532 | F8 | Group | 5.517 | 0.005 | 0.022 |
| Fp1 | Age | 1.746 | 0.189 | 0.225 | Fp1 | Group | 5.012 | 0.008 | 0.022 |
| Fp2 | Age | 0.565 | 0.454 | 0.507 | Fp2 | Group | 4.965 | 0.009 | 0.022 |
| Fz | Age | 4.244 | 0.042 | 0.066 | Fz | Group | 5.378 | 0.006 | 0.022 |
| O1 | Age | 5.463 | 0.021 | 0.065 | O1 | Group | 2.823 | 0.064 | 0.093 |
| O2 | Age | 5.245 | 0.024 | 0.065 | O2 | Group | 1.511 | 0.225 | 0.225 |
| P3 | Age | 4.474 | 0.037 | 0.066 | P3 | Group | 2.535 | 0.084 | 0.106 |
| P4 | Age | 4.299 | 0.041 | 0.066 | P4 | Group | 1.738 | 0.181 | 0.202 |
| P7 | Age | 1.930 | 0.168 | 0.212 | P7 | Group | 2.712 | 0.071 | 0.096 |
| P8 | Age | 3.251 | 0.074 | 0.101 | P8 | Group | 1.550 | 0.217 | 0.225 |
| Pz | Age | 6.100 | 0.015 | 0.060 | Pz | Group | 1.997 | 0.141 | 0.167 |
| T7 | Age | 6.886 | 0.010 | 0.060 | T7 | Group | 4.756 | 0.010 | 0.022 |
| T8 | Age | 0.393 | 0.532 | 0.532 | T8 | Group | 3.925 | 0.023 | 0.039 |
| C3 | edu | 0.046 | 0.830 | 0.995 | C3 | sex | 11.269 | 0.001 | 0.003 |
| C4 | edu | 0.019 | 0.890 | 0.995 | C4 | sex | 7.636 | 0.007 | 0.013 |
| Cz | edu | 0.122 | 0.728 | 0.995 | Cz | sex | 6.810 | 0.010 | 0.018 |
| F3 | edu | 0.368 | 0.545 | 0.995 | F3 | sex | 4.416 | 0.038 | 0.048 |
| F4 | edu | 0.531 | 0.468 | 0.995 | F4 | sex | 4.475 | 0.037 | 0.048 |
| F7 | edu | 0.937 | 0.335 | 0.995 | F7 | sex | 2.582 | 0.111 | 0.132 |
| F8 | edu | 0.193 | 0.661 | 0.995 | F8 | sex | 1.483 | 0.226 | 0.238 |
| Fp1 | edu | 0.287 | 0.593 | 0.995 | Fp1 | sex | 1.114 | 0.294 | 0.294 |
| Fp2 | edu | 0.267 | 0.607 | 0.995 | Fp2 | sex | 1.789 | 0.184 | 0.205 |
| Fz | edu | 0.697 | 0.406 | 0.995 | Fz | sex | 4.518 | 0.036 | 0.048 |
| O1 | edu | 0.009 | 0.924 | 0.995 | O1 | sex | 14.737 | 0.000 | 0.001 |
| O2 | edu | 0.011 | 0.915 | 0.995 | O2 | sex | 18.509 | 0.000 | 0.001 |
| P3 | edu | 0.037 | 0.847 | 0.995 | P3 | sex | 16.653 | 0.000 | 0.001 |
| P4 | edu | 0.106 | 0.745 | 0.995 | P4 | sex | 13.670 | 0.000 | 0.001 |
| P7 | edu | 0.732 | 0.394 | 0.995 | P7 | sex | 15.115 | 0.000 | 0.001 |
| P8 | edu | 0.109 | 0.742 | 0.995 | P8 | sex | 11.206 | 0.001 | 0.003 |
| Pz | edu | 0.003 | 0.960 | 0.995 | Pz | sex | 10.802 | 0.001 | 0.003 |
| T7 | edu | 0.286 | 0.594 | 0.995 | T7 | sex | 9.218 | 0.003 | 0.006 |
| T8 | edu | 0.000 | 0.995 | 0.995 | T8 | sex | 6.090 | 0.015 | 0.024 |

**Table S14.** Statistical values of the SLSW theta downward slope difference (Figure S3A) between groups (p-values from the linear mixed models), with amplitude as the dependent variable, and Group as the fixed effect. Age, sex, and education were used as covariates of no interest

*SLSW theta upward slope post-hoc comparisons*

| chan | effect | F | p | chan | effect | F | p | chan | effect | F | p |
|---|---|---|---|---|---|---|---|---|---|---|---|
| C3 | PD-H vs. PD-nH | 5.806 | 0.018 | C3 | PD-H vs. HC | 5.903 | 0.018 | C3 | PD-NH vs. HC | 0.123 | 0.727 |
| C4 | PD-H vs. PD-nH | 6.107 | 0.016 | C4 | PD-H vs. HC | 6.238 | 0.015 | C4 | PD-NH vs. HC | 0.036 | 0.851 |
| Cz | PD-H vs. PD-nH | 4.299 | 0.041 | Cz | PD-H vs. HC | 9.809 | 0.003 | Cz | PD-NH vs. HC | 0.751 | 0.389 |
| F3 | PD-H vs. PD-nH | 7.374 | 0.008 | F3 | PD-H vs. HC | 9.258 | 0.003 | F3 | PD-NH vs. HC | 0.529 | 0.470 |
| F4 | PD-H vs. PD-nH | 6.004 | 0.016 | F4 | PD-H vs. HC | 8.227 | 0.006 | F4 | PD-NH vs. HC | 0.343 | 0.560 |
| F7 | PD-H vs. PD-nH | 4.251 | 0.043 | F7 | PD-H vs. HC | 6.727 | 0.012 | F7 | PD-NH vs. HC | 0.351 | 0.555 |
| F8 | PD-H vs. PD-nH | 5.700 | 0.019 | F8 | PD-H vs. HC | 5.692 | 0.020 | F8 | PD-NH vs. HC | 0.075 | 0.786 |
| Fp1 | PD-H vs. PD-nH | 4.195 | 0.044 | Fp1 | PD-H vs. HC | 10.028 | 0.002 | Fp1 | PD-NH vs. HC | 0.961 | 0.330 |
| Fp2 | PD-H vs. PD-nH | 4.705 | 0.033 | Fp2 | PD-H vs. HC | 9.265 | 0.003 | Fp2 | PD-NH vs. HC | 0.607 | 0.438 |
| Fz | PD-H vs. PD-nH | 5.657 | 0.020 | Fz | PD-H vs. HC | 9.390 | 0.003 | Fz | PD-NH vs. HC | 0.569 | 0.453 |
| T7 | PD-H vs. PD-nH | 6.892 | 0.010 | T7 | PD-H vs. HC | 4.056 | 0.048 | T7 | PD-NH vs. HC | 0.001 | 0.980 |
| T8 | PD-H vs. PD-nH | 6.116 | 0.016 | T8 | PD-H vs. HC | 3.062 | 0.085 | T8 | PD-NH vs. HC | 0.009 | 0.925 |

**Table S15.** SLSW upward slope post-hoc comparisons on the significant clusters shown in Figure S3A. Here, in the linear mixed models, we also control for age, education, and sex.

*SLSW delta upward slope*

| chan | effect | F | p | pfdr | chan | effect | F | p | pfdr |
|---|---|---|---|---|---|---|---|---|---|
| C3 | Age | 5.157 | 0.025 | 0.111 | C3 | Group | 3.077 | 0.050 | 0.079 |
| C4 | Age | 4.585 | 0.034 | 0.111 | C4 | Group | 4.549 | 0.013 | 0.039 |
| Cz | Age | 6.401 | 0.013 | 0.111 | Cz | Group | 4.046 | 0.020 | 0.039 |
| F3 | Age | 4.654 | 0.033 | 0.111 | F3 | Group | 4.264 | 0.017 | 0.039 |
| F4 | Age | 3.333 | 0.071 | 0.122 | F4 | Group | 4.982 | 0.009 | 0.032 |
| F7 | Age | 3.653 | 0.059 | 0.111 | F7 | Group | 3.923 | 0.023 | 0.039 |
| F8 | Age | 0.180 | 0.672 | 0.672 | F8 | Group | 5.390 | 0.006 | 0.028 |
| Fp1 | Age | 1.692 | 0.196 | 0.266 | Fp1 | Group | 5.406 | 0.006 | 0.028 |
| Fp2 | Age | 0.774 | 0.381 | 0.426 | Fp2 | Group | 5.538 | 0.005 | 0.028 |
| Fz | Age | 3.789 | 0.054 | 0.111 | Fz | Group | 5.596 | 0.005 | 0.028 |
| O1 | Age | 3.006 | 0.086 | 0.136 | O1 | Group | 2.854 | 0.062 | 0.091 |
| O2 | Age | 4.028 | 0.047 | 0.111 | O2 | Group | 1.781 | 0.173 | 0.183 |
| P3 | Age | 2.628 | 0.108 | 0.158 | P3 | Group | 2.001 | 0.140 | 0.166 |
| P4 | Age | 3.862 | 0.052 | 0.111 | P4 | Group | 2.111 | 0.126 | 0.160 |
| P7 | Age | 1.389 | 0.241 | 0.286 | P7 | Group | 2.683 | 0.073 | 0.099 |
| P8 | Age | 1.522 | 0.220 | 0.279 | P8 | Group | 1.469 | 0.235 | 0.235 |
| Pz | Age | 4.180 | 0.043 | 0.111 | Pz | Group | 1.885 | 0.157 | 0.175 |
| T7 | Age | 3.946 | 0.050 | 0.111 | T7 | Group | 3.922 | 0.023 | 0.039 |
| T8 | Age | 0.239 | 0.626 | 0.660 | T8 | Group | 3.961 | 0.022 | 0.039 |
| C3 | edu | 0.236 | 0.628 | 0.972 | C3 | sex | 9.476 | 0.003 | 0.006 |
| C4 | edu | 0.001 | 0.972 | 0.972 | C4 | sex | 8.648 | 0.004 | 0.008 |
| Cz | edu | 0.022 | 0.883 | 0.972 | Cz | sex | 6.923 | 0.010 | 0.015 |
| F3 | edu | 0.539 | 0.465 | 0.972 | F3 | sex | 4.103 | 0.045 | 0.057 |
| F4 | edu | 0.194 | 0.661 | 0.972 | F4 | sex | 4.167 | 0.044 | 0.057 |
| F7 | edu | 0.564 | 0.454 | 0.972 | F7 | sex | 2.659 | 0.106 | 0.118 |
| F8 | edu | 0.024 | 0.876 | 0.972 | F8 | sex | 1.609 | 0.207 | 0.207 |
| Fp1 | edu | 0.358 | 0.551 | 0.972 | Fp1 | sex | 2.104 | 0.150 | 0.158 |
| Fp2 | edu | 0.148 | 0.701 | 0.972 | Fp2 | sex | 2.875 | 0.093 | 0.110 |
| Fz | edu | 0.320 | 0.573 | 0.972 | Fz | sex | 4.539 | 0.035 | 0.052 |
| O1 | edu | 0.463 | 0.498 | 0.972 | O1 | sex | 15.609 | 0.000 | 0.001 |
| O2 | edu | 0.148 | 0.701 | 0.972 | O2 | sex | 19.217 | 0.000 | 0.001 |
| P3 | edu | 0.037 | 0.848 | 0.972 | P3 | sex | 16.344 | 0.000 | 0.001 |
| P4 | edu | 0.014 | 0.905 | 0.972 | P4 | sex | 14.123 | 0.000 | 0.001 |
| P7 | edu | 0.651 | 0.422 | 0.972 | P7 | sex | 14.375 | 0.000 | 0.001 |
| P8 | edu | 0.586 | 0.446 | 0.972 | P8 | sex | 12.178 | 0.001 | 0.002 |
| Pz | edu | 0.002 | 0.965 | 0.972 | Pz | sex | 10.157 | 0.002 | 0.005 |
| T7 | edu | 0.142 | 0.707 | 0.972 | T7 | sex | 9.986 | 0.002 | 0.005 |
| T8 | edu | 0.175 | 0.677 | 0.972 | T8 | sex | 7.559 | 0.007 | 0.012 |

**Table S16.** Statistical values of the SLSW theta upward slope difference (Figure S3B) between groups (p-values from the linear mixed models), with amplitude as the dependent variable, and Group as the fixed effect. Age, sex, and education were used as covariates of no interest.

*SLSW delta upward slope post-hoc comparisons*

| chan | effect | F | p | chan | effect | F | p | chan | effect | F | p |
|---|---|---|---|---|---|---|---|---|---|---|---|
| C4 | PD-H vs. PD-nH | 6.841 | 0.011 | C4 | PD-H vs. HC | 6.932 | 0.011 | C4 | PD-nH vs. HC | 8.97E-05 | 0.992 |
| Cz | PD-H vs. PD-nH | 5.094 | 0.027 | Cz | PD-H vs. HC | 10.056 | 0.002 | Cz | PD-nH vs. HC | 0.258 | 0.613 |
| F3 | PD-H vs. PD-nH | 4.275 | 0.042 | F3 | PD-H vs. HC | 6.283 | 0.015 | F3 | PD-nH vs. HC | 0.328 | 0.569 |
| F4 | PD-H vs. PD-nH | 5.483 | 0.022 | F4 | PD-H vs. HC | 8.488 | 0.005 | F4 | PD-nH vs. HC | 0.424 | 0.517 |
| F7 | PD-H vs. PD-nH | 4.585 | 0.035 | F7 | PD-H vs. HC | 6.393 | 0.014 | F7 | PD-nH vs. HC | 0.415 | 0.521 |
| F8 | PD-H vs. PD-nH | 7.826 | 0.006 | F8 | PD-H vs. HC | 5.976 | 0.017 | F8 | PD-nH vs. HC | 0.005 | 0.947 |
| Fp1 | PD-H vs. PD-nH | 4.286 | 0.042 | Fp1 | PD-H vs. HC | 9.447 | 0.003 | Fp1 | PD-nH vs. HC | 1.218 | 0.273 |
| Fp2 | PD-H vs. PD-nH | 5.977 | 0.017 | Fp2 | PD-H vs. HC | 7.988 | 0.006 | Fp2 | PD-nH vs. HC | 0.262 | 0.610 |
| Fz | PD-H vs. PD-nH | 5.257 | 0.025 | Fz | PD-H vs. HC | 10.490 | 0.002 | Fz | PD-nH vs. HC | 0.920 | 0.341 |
| T7 | PD-H vs. PD-nH | 6.609 | 0.012 | T7 | PD-H vs. HC | 2.874 | 0.095 | T7 | PD-nH vs. HC | 0.130 | 0.720 |
| T8 | PD-H vs. PD-nH | 6.067 | 0.016 | T8 | PD-H vs. HC | 2.812 | 0.099 | T8 | PD-nH vs. HC | 0.023 | 0.879 |

**Table S17.** SLSW upward slope post-hoc comparisons on the significant clusters shown in Figure S3A. Here, in the linear mixed models, we also control for age, education, and sex.

*The cumulative sum of peak-to-peak amplitude of theta SLSW as an index of PD and PD hallucinations*

| chan | effect | F | p | pfdr | chan | effect | F | p | pfdr |
|---|---|---|---|---|---|---|---|---|---|
| C3 | Age | 1.92 | 0.163 | 0.417 | C3 | Group | 9.261 | 0 | 0.002 |
| C4 | Age | 2.337 | 0.134 | 0.417 | C4 | Group | 5.746 | 0.005 | 0.01 |
| Cz | Age | 1.135 | 0.284 | 0.491 | Cz | Group | 6.667 | 0.002 | 0.005 |
| F3 | Age | 0.777 | 0.385 | 0.538 | F3 | Group | 9.68 | 0 | 0.002 |
| F4 | Age | 1.4 | 0.243 | 0.471 | F4 | Group | 6.824 | 0.002 | 0.005 |
| F7 | Age | 0.308 | 0.568 | 0.674 | F7 | Group | 6.78 | 0.001 | 0.005 |
| F8 | Age | 0.113 | 0.744 | 0.744 | F8 | Group | 4.642 | 0.012 | 0.023 |
| Fp1 | Age | 0.21 | 0.651 | 0.687 | Fp1 | Group | 7.771 | 0 | 0.002 |
| Fp2 | Age | 0.394 | 0.537 | 0.674 | Fp2 | Group | 6.929 | 0.002 | 0.005 |
| Fz | Age | 1.322 | 0.248 | 0.471 | Fz | Group | 9.127 | 0 | 0.002 |
| O1 | Age | 2.732 | 0.097 | 0.417 | O1 | Group | 2.307 | 0.112 | 0.126 |
| O2 | Age | 3.671 | 0.056 | 0.417 | O2 | Group | 2.056 | 0.126 | 0.133 |
| P3 | Age | 2.154 | 0.142 | 0.417 | P3 | Group | 4.324 | 0.015 | 0.026 |
| P4 | Age | 1.894 | 0.176 | 0.417 | P4 | Group | 2.402 | 0.096 | 0.121 |
| P7 | Age | 0.984 | 0.331 | 0.524 | P7 | Group | 4.003 | 0.02 | 0.029 |
| P8 | Age | 0.743 | 0.396 | 0.538 | P8 | Group | 1.721 | 0.185 | 0.185 |
| Pz | Age | 2.231 | 0.136 | 0.417 | Pz | Group | 3.629 | 0.028 | 0.039 |
| T7 | Age | 2.342 | 0.129 | 0.417 | T7 | Group | 4.098 | 0.017 | 0.028 |
| T8 | Age | 0.25 | 0.615 | 0.687 | T8 | Group | 2.301 | 0.104 | 0.124 |
| C3 | education | 0 | 0.987 | 0.993 | C3 | sex | 4.311 | 0.042 | 0.057 |
| C4 | education | 0.055 | 0.81 | 0.993 | C4 | sex | 5.546 | 0.019 | 0.04 |
| Cz | education | 0.013 | 0.909 | 0.993 | Cz | sex | 2.44 | 0.119 | 0.124 |
| F3 | education | 0.014 | 0.909 | 0.993 | F3 | sex | 4.655 | 0.03 | 0.057 |
| F4 | education | 0.007 | 0.933 | 0.993 | F4 | sex | 4.138 | 0.045 | 0.057 |
| F7 | education | 0.128 | 0.725 | 0.993 | F7 | sex | 2.404 | 0.124 | 0.124 |
| F8 | education | 0.053 | 0.821 | 0.993 | F8 | sex | 2.716 | 0.103 | 0.115 |
| Fp1 | education | 0 | 0.984 | 0.993 | Fp1 | sex | 2.73 | 0.1 | 0.115 |
| Fp2 | education | 0.021 | 0.885 | 0.993 | Fp2 | sex | 4.264 | 0.044 | 0.057 |
| Fz | education | 0 | 0.993 | 0.993 | Fz | sex | 4.376 | 0.036 | 0.057 |
| O1 | education | 0 | 0.986 | 0.993 | O1 | sex | 11.48 | 0.001 | 0.01 |
| O2 | education | 0.18 | 0.676 | 0.993 | O2 | sex | 12.61 | 0.001 | 0.01 |
| P3 | education | 0.066 | 0.788 | 0.993 | P3 | sex | 8.266 | 0.004 | 0.013 |
| P4 | education | 0.06 | 0.801 | 0.993 | P4 | sex | 8.73 | 0.003 | 0.013 |
| P7 | education | 0.007 | 0.94 | 0.993 | P7 | sex | 10.311 | 0.002 | 0.013 |
| P8 | education | 0.172 | 0.684 | 0.993 | P8 | sex | 9.24 | 0.003 | 0.013 |
| Pz | education | 0.002 | 0.965 | 0.993 | Pz | sex | 5.762 | 0.017 | 0.04 |
| T7 | education | 1.226 | 0.274 | 0.993 | T7 | sex | 4.333 | 0.041 | 0.057 |
| T8 | education | 0.171 | 0.678 | 0.993 | T8 | sex | 6.169 | 0.015 | 0.04 |

**Table S18.** Statistical values of the SLSW theta cumulative sum of the peak-to-peak amplitude (permutation p-values from the ANCOVA), with density as the dependent variable, and Group as the independent variable. Age, sex, and education were used as covariates of no interest.

*Post-hoc comparisons of the cumulative sum of peak-to-peak amplitude of theta SLSW*

| chan | effect | F | p | chan | effect | F | p | chan | effect | F | p |
|---|---|---|---|---|---|---|---|---|---|---|---|
| C3 | PD-H vs. PD-nH | 7.269 | 0.008 | C3 | PD-H vs. HC | 14.507 | 0 | C3 | PD-nH vs. HC | 4.239 | 0.042 |
| C4 | PD-H vs. PD-nH | 4.449 | 0.036 | C4 | PD-H vs. HC | 9.018 | 0.003 | C4 | PD-nH vs. HC | 2.391 | 0.128 |
| Cz | PD-H vs. PD-nH | 3.942 | 0.048 | Cz | PD-H vs. HC | 11.833 | 0.002 | Cz | PD-nH vs. HC | 3.849 | 0.052 |
| F3 | PD-H vs. PD-nH | 7.687 | 0.007 | F3 | PD-H vs. HC | 15.229 | 0 | F3 | PD-nH vs. HC | 4.009 | 0.046 |
| F4 | PD-H vs. PD-nH | 4.597 | 0.032 | F4 | PD-H vs. HC | 10.711 | 0.002 | F4 | PD-nH vs. HC | 4.248 | 0.041 |
| F7 | PD-H vs. PD-nH | 5.426 | 0.02 | F7 | PD-H vs. HC | 9.233 | 0.003 | F7 | PD-nH vs. HC | 3.496 | 0.067 |
| F8 | PD-H vs. PD-nH | 3.301 | 0.07 | F8 | PD-H vs. HC | 6.973 | 0.01 | F8 | PD-nH vs. HC | 2.934 | 0.092 |
| Fp1 | PD-H vs. PD-nH | 5.357 | 0.024 | Fp1 | PD-H vs. HC | 12.456 | 0.001 | Fp1 | PD-nH vs. HC | 4.152 | 0.042 |
| Fp2 | PD-H vs. PD-nH | 4.895 | 0.028 | Fp2 | PD-H vs. HC | 10.993 | 0.001 | Fp2 | PD-nH vs. HC | 3.994 | 0.051 |
| Fz | PD-H vs. PD-nH | 6.79 | 0.01 | Fz | PD-H vs. HC | 14.403 | 0 | Fz | PD-nH vs. HC | 4.158 | 0.042 |
| P3 | PD-H vs. PD-nH | 3.383 | 0.069 | P3 | PD-H vs. HC | 7.63 | 0.005 | P3 | PD-nH vs. HC | 1.406 | 0.246 |
| P7 | PD-H vs. PD-nH | 4.069 | 0.047 | P7 | PD-H vs. HC | 5.349 | 0.022 | P7 | PD-nH vs. HC | 0.855 | 0.367 |
| Pz | PD-H vs. PD-nH | 2.254 | 0.134 | Pz | PD-H vs. HC | 7.888 | 0.007 | Pz | PD-nH vs. HC | 1.473 | 0.225 |
| T7 | PD-H vs. PD-nH | 2.274 | 0.136 | T7 | PD-H vs. HC | 5.998 | 0.016 | T7 | PD-nH vs. HC | 3.302 | 0.072 |

**Table S19.** SLSW cumulative sum of peak-to-peak amplitude comparisons on the significant clusters shown in Figure 3 and Table S18. Here, in the linear mixed models, we also control for age, education, and sex.

*The cumulative sum of peak-to-peak amplitude of delta SLSW as an index of PD and PD hallucinations*

| chan | effect | F | p | pfdr | chan | effect | F | p | pfdr |
|---|---|---|---|---|---|---|---|---|---|
| C3 | Age | 11.77 | 0 | 0 | C3 | Group | 1.95 | 0.15 | 0.15 |
| C4 | Age | 7.88 | 0.01 | 0.032 | C4 | Group | 4.95 | 0.01 | 0.032 |
| Cz | Age | 12.75 | 0 | 0 | Cz | Group | 2.79 | 0.07 | 0.083 |
| F3 | Age | 10.03 | 0 | 0 | F3 | Group | 3.17 | 0.05 | 0.063 |
| F4 | Age | 8.87 | 0 | 0 | F4 | Group | 3.49 | 0.03 | 0.047 |
| F7 | Age | 5.04 | 0.03 | 0.071 | F7 | Group | 3.69 | 0.03 | 0.047 |
| F8 | Age | 0.37 | 0.54 | 0.54 | F8 | Group | 5.42 | 0.01 | 0.032 |
| Fp1 | Age | 2.45 | 0.12 | 0.19 | Fp1 | Group | 2.28 | 0.11 | 0.116 |
| Fp2 | Age | 0.82 | 0.37 | 0.414 | Fp2 | Group | 2.54 | 0.08 | 0.089 |
| Fz | Age | 10.62 | 0 | 0 | Fz | Group | 3.5 | 0.03 | 0.047 |
| O1 | Age | 0.44 | 0.5 | 0.528 | O1 | Group | 6.54 | 0 | 0 |
| O2 | Age | 1.02 | 0.31 | 0.405 | O2 | Group | 5.61 | 0 | 0 |
| P3 | Age | 3.95 | 0.05 | 0.095 | P3 | Group | 3.54 | 0.03 | 0.047 |
| P4 | Age | 3 | 0.09 | 0.155 | P4 | Group | 4.81 | 0.01 | 0.032 |
| P7 | Age | 0.86 | 0.35 | 0.414 | P7 | Group | 4.29 | 0.02 | 0.047 |
| P8 | Age | 1.71 | 0.2 | 0.292 | P8 | Group | 3.8 | 0.02 | 0.047 |
| Pz | Age | 4.28 | 0.05 | 0.095 | Pz | Group | 3.06 | 0.05 | 0.063 |
| T7 | Age | 5.69 | 0.02 | 0.054 | T7 | Group | 3.15 | 0.05 | 0.063 |
| T8 | Age | 1.05 | 0.32 | 0.405 | T8 | Group | 4.85 | 0.01 | 0.032 |
| C3 | education | 2.76 | 0.1 | 0.38 | C3 | sex | 8.92 | 0 | 0 |
| C4 | education | 0.77 | 0.38 | 0.602 | C4 | sex | 9.31 | 0 | 0 |
| Cz | education | 2.44 | 0.12 | 0.38 | Cz | sex | 7.27 | 0.01 | 0.015 |
| F3 | education | 3.03 | 0.08 | 0.38 | F3 | sex | 3.45 | 0.07 | 0.089 |
| F4 | education | 2.56 | 0.11 | 0.38 | F4 | sex | 6.28 | 0.01 | 0.015 |
| F7 | education | 2.57 | 0.11 | 0.38 | F7 | sex | 2.81 | 0.1 | 0.119 |
| F8 | education | 1.49 | 0.23 | 0.475 | F8 | sex | 0.41 | 0.52 | 0.52 |
| Fp1 | education | 1.39 | 0.25 | 0.475 | Fp1 | sex | 2.03 | 0.16 | 0.169 |
| Fp2 | education | 0.94 | 0.33 | 0.57 | Fp2 | sex | 2.01 | 0.16 | 0.169 |
| Fz | education | 2.33 | 0.13 | 0.38 | Fz | sex | 6.22 | 0.01 | 0.015 |
| O1 | education | 2.15 | 0.14 | 0.38 | O1 | sex | 14.46 | 0 | 0 |
| O2 | education | 0.54 | 0.47 | 0.687 | O2 | sex | 20.58 | 0 | 0 |
| P3 | education | 0.15 | 0.7 | 0.816 | P3 | sex | 17.11 | 0 | 0 |
| P4 | education | 0.04 | 0.83 | 0.876 | P4 | sex | 11.17 | 0 | 0 |
| P7 | education | 0.22 | 0.64 | 0.811 | P7 | sex | 14.6 | 0 | 0 |
| P8 | education | 0.13 | 0.73 | 0.816 | P8 | sex | 11.29 | 0 | 0 |
| Pz | education | 0.02 | 0.88 | 0.88 | Pz | sex | 9.66 | 0 | 0 |
| T7 | education | 1.57 | 0.22 | 0.475 | T7 | sex | 7.72 | 0 | 0 |
| T8 | education | 0.31 | 0.59 | 0.801 | T8 | sex | 3.63 | 0.06 | 0.081 |

**Table S20.** Statistical values of the SLSW theta cumulative sum of the peak-to-peak amplitude, permutation p-values from the ANCOVA, with density as the dependent variable, and Group as the independent variable. Age, sex, and education were used as covariates of no interest.

*Post-hoc comparisons of the cumulative sum of peak-to-peak amplitude of delta SLSW*

| chan | effect | F | p | chan | effect | F | p | chan | effect | F | p |
|---|---|---|---|---|---|---|---|---|---|---|---|
| C4 | PD-H vs. PD-nH | 6.41 | 0.012 | C4 | PD-H vs. HC | 6.825 | 0.01 | C4 | PD-nH vs. HC | 0.239 | 0.626 |
| F8 | PD-H vs. PD-nH | 5.778 | 0.02 | F8 | PD-H vs. HC | 5.921 | 0.016 | F8 | PD-nH vs. HC | 2.227 | 0.142 |
| O1 | PD-H vs. PD-nH | 6.572 | 0.012 | O1 | PD-H vs. HC | 8.45 | 0.005 | O1 | PD-nH vs. HC | 1.618 | 0.206 |
| O2 | PD-H vs. PD-nH | 5.53 | 0.021 | O2 | PD-H vs. HC | 7.824 | 0.005 | O2 | PD-nH vs. HC | 1.884 | 0.181 |
| P4 | PD-H vs. PD-nH | 3.586 | 0.062 | P4 | PD-H vs. HC | 7.777 | 0.007 | P4 | PD-nH vs. HC | 2.347 | 0.127 |
| P7 | PD-H vs. PD-nH | 4.278 | 0.039 | P7 | PD-H vs. HC | 5.348 | 0.024 | P7 | PD-nH vs. HC | 1.285 | 0.27 |
| T8 | PD-H vs. PD-nH | 6.856 | 0.01 | T8 | PD-H vs. HC | 4.154 | 0.048 | T8 | PD-nH vs. HC | 0.635 | 0.43 |

**Table S21.** SLSW cumulative sum of peak-to-peak amplitude comparisons on the significant clusters shown in Figure 3 and Table S18. Here, in the linear mixed models, we also control for age, education, and sex.

*SLSW theta peak-to-peak amplitude association with the severity of the hallucinatory burden*

| chan | estimate | F | p | pfdr |
|---|---|---|---|---|
| C3 | 0.073 | 2.434 | 0.017 | 0.047 |
| C4 | 0.068 | 2.439 | 0.017 | 0.047 |
| Cz | 0.062 | 2.150 | 0.035 | 0.079 |
| F3 | 0.077 | 2.685 | 0.009 | 0.043 |
| F4 | 0.071 | 2.590 | 0.011 | 0.043 |
| F7 | 0.075 | 2.115 | 0.038 | 0.079 |
| F8 | 0.058 | 1.824 | 0.072 | 0.098 |
| Fp1 | 0.057 | 1.887 | 0.063 | 0.098 |
| Fp2 | 0.055 | 1.847 | 0.069 | 0.098 |
| Fz | 0.067 | 2.611 | 0.011 | 0.043 |
| O1 | 0.055 | 1.706 | 0.092 | 0.109 |
| O2 | 0.040 | 1.280 | 0.204 | 0.204 |
| P3 | 0.059 | 2.058 | 0.043 | 0.082 |
| P4 | 0.042 | 1.382 | 0.171 | 0.180 |
| P7 | 0.075 | 2.626 | 0.010 | 0.043 |
| P8 | 0.055 | 1.956 | 0.054 | 0.093 |
| Pz | 0.043 | 1.469 | 0.146 | 0.163 |
| T7 | 0.096 | 3.148 | 0.002 | 0.043 |
| T8 | 0.054 | 1.762 | 0.082 | 0.104 |

**Table S22.** Higher SLSW theta peak-to-peak amplitude is associated with a more severe hallucinatory burden.

*SLSW delta peak-to-peak amplitude association with the severity of the hallucinatory burden*

| chan | estimate | F | p | pfdr |
|---|---|---|---|---|
| C3 | 0.050 | 2.124 | 0.037 | 0.107 |
| C4 | 0.048 | 2.225 | 0.029 | 0.107 |
| Cz | 0.041 | 1.891 | 0.062 | 0.107 |
| F3 | 0.044 | 1.898 | 0.061 | 0.107 |
| F4 | 0.042 | 2.073 | 0.041 | 0.107 |
| F7 | 0.043 | 1.608 | 0.112 | 0.133 |
| F8 | 0.031 | 1.429 | 0.157 | 0.157 |
| Fp1 | 0.034 | 1.509 | 0.135 | 0.148 |
| Fp2 | 0.031 | 1.491 | 0.140 | 0.148 |
| Fz | 0.042 | 2.131 | 0.036 | 0.107 |
| O1 | 0.050 | 2.290 | 0.025 | 0.107 |
| O2 | 0.041 | 1.889 | 0.063 | 0.107 |
| P3 | 0.040 | 1.991 | 0.050 | 0.107 |
| P4 | 0.036 | 1.701 | 0.093 | 0.129 |
| P7 | 0.045 | 2.248 | 0.027 | 0.107 |
| P8 | 0.035 | 1.854 | 0.067 | 0.107 |
| Pz | 0.036 | 1.649 | 0.103 | 0.131 |
| T7 | 0.047 | 2.393 | 0.019 | 0.107 |
| T8 | 0.035 | 1.689 | 0.095 | 0.129 |

**Table S23.** Higher SLSW delta peak-to-peak amplitude is associated with a more severe hallucinatory burden, but no significant difference was observed after FDR correction.

*SLSW theta cumulative sum of the peak-to-peak amplitude association with the severity of the hallucinatory burden*

| chan | estimate | F | p | pfdr |
|---|---|---|---|---|
| C3 | 13.850 | 3.481 | 0.001 | 0.004 |
| C4 | 12.386 | 3.062 | 0.003 | 0.006 |
| Cz | 11.505 | 2.972 | 0.004 | 0.007 |
| F3 | 13.790 | 3.670 | 0.000 | 0.004 |
| F4 | 12.736 | 3.276 | 0.002 | 0.004 |
| F7 | 15.377 | 2.990 | 0.004 | 0.007 |
| F8 | 13.687 | 2.715 | 0.008 | 0.013 |
| Fp1 | 16.242 | 3.443 | 0.001 | 0.004 |
| Fp2 | 15.446 | 3.427 | 0.001 | 0.004 |
| Fz | 12.822 | 3.642 | 0.000 | 0.004 |
| O1 | 12.127 | 1.961 | 0.053 | 0.066 |
| O2 | 6.554 | 1.077 | 0.285 | 0.285 |
| P3 | 12.191 | 2.606 | 0.011 | 0.016 |
| P4 | 9.487 | 1.916 | 0.059 | 0.066 |
| P7 | 16.962 | 3.304 | 0.001 | 0.004 |
| P8 | 12.262 | 2.387 | 0.019 | 0.026 |
| Pz | 9.464 | 1.936 | 0.056 | 0.066 |
| T7 | 18.471 | 3.077 | 0.003 | 0.006 |
| T8 | 10.230 | 1.834 | 0.070 | 0.074 |

**Table S24.** Higher SLSW theta cumulative sum of the peak-to-peak amplitude is associated with a more severe hallucinatory burden, but no significant difference was observed after FDR correction.

*SLSW delta cumulative sum of the peak-to-peak amplitude association with the severity of the hallucinatory burden*

| chan | estimate | F | p | pfdr |
|---|---|---|---|---|
| C3 | 4.586 | 1.196 | 0.235 | 0.342 |
| C4 | 6.465 | 2.023 | 0.046 | 0.182 |
| Cz | 4.910 | 1.586 | 0.117 | 0.230 |
| F3 | 4.201 | 1.246 | 0.217 | 0.342 |
| F4 | 3.534 | 1.147 | 0.255 | 0.342 |
| F7 | 4.123 | 1.002 | 0.320 | 0.380 |
| F8 | 2.980 | 0.874 | 0.385 | 0.430 |
| Fp1 | 2.337 | 0.707 | 0.482 | 0.509 |
| Fp2 | 1.196 | 0.360 | 0.720 | 0.720 |
| Fz | 3.110 | 1.111 | 0.270 | 0.342 |
| O1 | 8.148 | 2.386 | 0.019 | 0.182 |
| O2 | 8.008 | 2.640 | 0.010 | 0.182 |
| P3 | 6.524 | 1.879 | 0.064 | 0.202 |
| P4 | 5.427 | 1.698 | 0.093 | 0.222 |
| P7 | 7.769 | 2.009 | 0.048 | 0.182 |
| P8 | 7.067 | 2.092 | 0.040 | 0.182 |
| Pz | 4.639 | 1.559 | 0.123 | 0.230 |
| T7 | 5.885 | 1.749 | 0.084 | 0.222 |
| T8 | 5.132 | 1.517 | 0.133 | 0.230 |

**Table S25.** Higher SLSW delta cumulative sum of the peak-to-peak amplitude is associated with a more severe hallucinatory burden, but no significant difference was observed after FDR correction.

*SLSW theta density is associated with cognitive impairment*

| chan | effect | F | p | pfdr | chan | effect | F | p | pfdr |
|---|---|---|---|---|---|---|---|---|---|
| C3 | age | -3.695 | 0.001 | 0.001 | C3 | edu | 3.604 | <0.001 | 0.001 |
| C4 | age | -3.954 | <0.001 | <0.001 | C4 | edu | 3.637 | 0.001 | 0.001 |
| Cz | age | -3.731 | <0.001 | <0.001 | Cz | edu | 3.795 | <0.001 | 0.001 |
| F3 | age | -3.577 | <0.001 | <0.001 | F3 | edu | 3.709 | <0.001 | 0.001 |
| F4 | age | -3.922 | <0.001 | <0.001 | F4 | edu | 3.76 | <0.001 | 0.001 |
| F7 | age | -3.618 | 0.001 | 0.001 | F7 | edu | 3.482 | 0.001 | 0.001 |
| F8 | age | -3.854 | <0.001 | <0.001 | F8 | edu | 3.568 | 0.001 | 0.001 |
| Fp1 | age | -3.695 | <0.001 | <0.001 | Fp1 | edu | 3.554 | <0.001 | 0.001 |
| Fp2 | age | -3.902 | <0.001 | <0.001 | Fp2 | edu | 3.538 | 0.001 | 0.001 |
| Fz | age | -3.909 | <0.001 | <0.001 | Fz | edu | 3.81 | <0.001 | 0.001 |
| O1 | age | -3.828 | <0.001 | <0.001 | O1 | edu | 3.363 | 0.002 | 0.002 |
| O2 | age | -3.893 | <0.001 | <0.001 | O2 | edu | 3.486 | <0.001 | 0.001 |
| P3 | age | -3.916 | <0.001 | <0.001 | P3 | edu | 3.562 | <0.001 | 0.001 |
| P4 | age | -3.913 | <0.001 | <0.001 | P4 | edu | 3.592 | 0.001 | 0.001 |
| P7 | age | -3.868 | <0.001 | <0.001 | P7 | edu | 3.33 | 0.002 | 0.002 |
| P8 | age | -3.81 | <0.001 | <0.001 | P8 | edu | 3.516 | 0.001 | 0.001 |
| Pz | age | -3.82 | <0.001 | <0.001 | Pz | edu | 3.551 | 0.001 | 0.001 |
| T7 | age | -3.816 | <0.001 | <0.001 | T7 | edu | 3.344 | 0.002 | 0.002 |
| T8 | age | -3.896 | <0.001 | <0.001 | T8 | edu | 3.411 | 0.001 | 0.001 |
| C3 | sex | 0.535 | 0.586 | 0.586 | C3 | density | -2.884 | 0.005 | 0.012 |
| C4 | sex | 1.021 | 0.309 | 0.497 | C4 | density | -3.809 | <0.001 | 0.001 |
| Cz | sex | 0.561 | 0.576 | 0.586 | Cz | density | -4.134 | <0.001 | 0.001 |
| F3 | sex | 0.997 | 0.314 | 0.497 | F3 | density | -3.024 | 0.003 | 0.01 |
| F4 | sex | 1.031 | 0.311 | 0.497 | F4 | density | -3.95 | <0.001 | 0.002 |
| F7 | sex | 0.909 | 0.366 | 0.497 | F7 | density | -2.042 | 0.044 | 0.065 |
| F8 | sex | 1.07 | 0.291 | 0.497 | F8 | density | -2.696 | 0.007 | 0.013 |
| Fp1 | sex | 1.015 | 0.308 | 0.497 | Fp1 | density | -2.714 | 0.007 | 0.013 |
| Fp2 | sex | 1.049 | 0.293 | 0.497 | Fp2 | density | -2.838 | 0.004 | 0.011 |
| Fz | sex | 0.94 | 0.344 | 0.497 | Fz | density | -4.421 | <0.001 | 0.001 |
| O1 | sex | 0.876 | 0.375 | 0.497 | O1 | density | -1.777 | 0.077 | 0.104 |
| O2 | sex | 0.91 | 0.36 | 0.497 | O2 | density | -1.635 | 0.106 | 0.134 |
| P3 | sex | 0.706 | 0.481 | 0.538 | P3 | density | -2.532 | 0.014 | 0.023 |
| P4 | sex | 0.923 | 0.361 | 0.497 | P4 | density | -2.778 | 0.007 | 0.013 |
| P7 | sex | 0.882 | 0.378 | 0.497 | P7 | density | -0.963 | 0.331 | 0.331 |
| P8 | sex | 0.943 | 0.349 | 0.497 | P8 | density | -1.1 | 0.276 | 0.291 |
| Pz | sex | 0.816 | 0.418 | 0.497 | Pz | density | -3.356 | 0.001 | 0.004 |
| T7 | sex | 0.821 | 0.409 | 0.497 | T7 | density | -1.238 | 0.21 | 0.235 |
| T8 | sex | 1.063 | 0.287 | 0.497 | T8 | density | -1.398 | 0.169 | 0.2 |

**Table S27.** Linear regression between theta SLSW density and cognitive PD-CRS score. Here, the linear regression controls age, education, and sex, with PD-CRS cognitive score as the dependent variable, and the indicated SLSW feature as the independent variable.

*SLSW delta density is not associated with cognitive impairment*

| chan | effect | F | p | pfdr | chan | effect | F | p | pfdr |
|---|---|---|---|---|---|---|---|---|---|
| C3 | age | -3.746 | <0.001 | <0.001 | C3 | edu | 3.497 | 0.001 | 0.001 |
| C4 | age | -3.832 | <0.001 | <0.001 | C4 | edu | 3.452 | 0.001 | 0.001 |
| Cz | age | -3.773 | <0.001 | <0.001 | Cz | edu | 3.462 | 0.001 | 0.001 |
| F3 | age | -3.822 | <0.001 | <0.001 | F3 | edu | 3.466 | 0.001 | 0.001 |
| F4 | age | -3.839 | <0.001 | <0.001 | F4 | edu | 3.435 | 0.001 | 0.002 |
| F7 | age | -3.907 | <0.001 | <0.001 | F7 | edu | 3.351 | 0.001 | 0.002 |
| F8 | age | -3.884 | <0.001 | <0.001 | F8 | edu | 3.35 | 0.001 | 0.001 |
| Fp1 | age | -3.85 | <0.001 | <0.001 | Fp1 | edu | 3.461 | 0.001 | 0.001 |
| Fp2 | age | -3.893 | <0.001 | <0.001 | Fp2 | edu | 3.437 | 0.001 | 0.001 |
| Fz | age | -3.822 | <0.001 | <0.001 | Fz | edu | 3.453 | 0.001 | 0.001 |
| O1 | age | -3.794 | <0.001 | 0.001 | O1 | edu | 3.504 | 0.001 | 0.001 |
| O2 | age | -3.805 | 0.001 | 0.001 | O2 | edu | 3.449 | 0.001 | 0.001 |
| P3 | age | -3.88 | <0.001 | <0.001 | P3 | edu | 3.49 | 0.001 | 0.001 |
| P4 | age | -3.853 | <0.001 | <0.001 | P4 | edu | 3.45 | 0.001 | 0.001 |
| P7 | age | -3.852 | <0.001 | <0.001 | P7 | edu | 3.403 | 0.002 | 0.002 |
| P8 | age | -3.894 | <0.001 | <0.001 | P8 | edu | 3.278 | 0.002 | 0.002 |
| Pz | age | -3.85 | <0.001 | <0.001 | Pz | edu | 3.487 | 0.001 | 0.001 |
| T7 | age | -3.879 | <0.001 | 0.001 | T7 | edu | 3.405 | 0.001 | 0.001 |
| T8 | age | -3.959 | <0.001 | <0.001 | T8 | edu | 3.336 | 0.002 | 0.002 |
| C3 | sex | 0.846 | 0.401 | 0.401 | C3 | density | 0.32 | 0.748 | 0.916 |
| C4 | sex | 0.898 | 0.372 | 0.4 | C4 | density | -0.192 | 0.852 | 0.953 |
| Cz | sex | 0.873 | 0.379 | 0.4 | Cz | density | 0.013 | 0.99 | 0.99 |
| F3 | sex | 0.883 | 0.378 | 0.4 | F3 | density | -0.063 | 0.949 | 0.99 |
| F4 | sex | 0.915 | 0.363 | 0.4 | F4 | density | -0.289 | 0.771 | 0.916 |
| F7 | sex | 0.982 | 0.324 | 0.4 | F7 | density | -1.296 | 0.198 | 0.873 |
| F8 | sex | 0.926 | 0.346 | 0.4 | F8 | density | -0.973 | 0.336 | 0.873 |
| Fp1 | sex | 0.918 | 0.366 | 0.4 | Fp1 | density | -0.344 | 0.732 | 0.916 |
| Fp2 | sex | 0.968 | 0.335 | 0.4 | Fp2 | density | -0.814 | 0.425 | 0.873 |
| Fz | sex | 0.918 | 0.362 | 0.4 | Fz | density | -0.3 | 0.757 | 0.916 |
| O1 | sex | 0.961 | 0.333 | 0.4 | O1 | density | -0.943 | 0.348 | 0.873 |
| O2 | sex | 1.004 | 0.32 | 0.4 | O2 | density | -1.187 | 0.232 | 0.873 |
| P3 | sex | 0.951 | 0.343 | 0.4 | P3 | density | -0.676 | 0.506 | 0.873 |
| P4 | sex | 0.934 | 0.355 | 0.4 | P4 | density | -0.718 | 0.473 | 0.873 |
| P7 | sex | 0.945 | 0.348 | 0.4 | P7 | density | -0.742 | 0.465 | 0.873 |
| P8 | sex | 1.065 | 0.291 | 0.4 | P8 | density | -1.621 | 0.106 | 0.873 |
| Pz | sex | 0.918 | 0.355 | 0.4 | Pz | density | -0.4 | 0.687 | 0.916 |
| T7 | sex | 0.931 | 0.354 | 0.4 | T7 | density | -0.774 | 0.435 | 0.873 |
| T8 | sex | 0.981 | 0.335 | 0.4 | T8 | density | -1.218 | 0.222 | 0.873 |

**Table S28.** Linear regression between delta SLSW density and cognitive PD-CRS score. Here, the linear regression controls age, education, and sex, with PD-CRS cognitive score as the dependent variable, and the indicated SLSW feature as the independent variable. No significant result was observed.

*SLSW theta peak-to-peak amplitude is associated with cognitive impairment*

| chan | effect | F | p | pfdr | chan | effect | F | p | pfdr |
|---|---|---|---|---|---|---|---|---|---|
| C3 | age | -4.285 | <0.001 | 0 | C3 | edu | 3.471 | 0.001 | 0.001 |
| C4 | age | -4.278 | <0.001 | 0 | C4 | edu | 3.517 | <0.001 | 0.001 |
| Cz | age | -4.174 | <0.001 | 0 | Cz | edu | 3.446 | 0.001 | 0.001 |
| F3 | age | -4.306 | <0.001 | 0 | F3 | edu | 3.39 | 0.001 | 0.001 |
| F4 | age | -4.237 | <0.001 | 0 | F4 | edu | 3.401 | 0.001 | 0.001 |
| F7 | age | -4.246 | <0.001 | 0 | F7 | edu | 3.319 | 0.001 | 0.001 |
| F8 | age | -3.904 | <0.001 | 0 | F8 | edu | 3.466 | 0.001 | 0.001 |
| Fp1 | age | -4.08 | <0.001 | 0 | Fp1 | edu | 3.42 | 0.001 | 0.001 |
| Fp2 | age | -3.98 | <0.001 | 0 | Fp2 | edu | 3.449 | 0.001 | 0.001 |
| Fz | age | -4.272 | <0.001 | 0 | Fz | edu | 3.398 | 0.001 | 0.001 |
| O1 | age | -3.953 | <0.001 | 0 | O1 | edu | 3.519 | 0.001 | 0.001 |
| O2 | age | -4.089 | <0.001 | 0 | O2 | edu | 3.553 | 0.001 | 0.001 |
| P3 | age | -4.168 | <0.001 | 0 | P3 | edu | 3.544 | 0.002 | 0.002 |
| P4 | age | -4.129 | <0.001 | 0 | P4 | edu | 3.544 | 0.001 | 0.001 |
| P7 | age | -3.976 | <0.001 | 0 | P7 | edu | 3.639 | <0.001 | 0.001 |
| P8 | age | -4.072 | <0.001 | 0 | P8 | edu | 3.576 | 0.001 | 0.001 |
| Pz | age | -4.17 | <0.001 | 0 | Pz | edu | 3.5 | 0.001 | 0.001 |
| T7 | age | -4.094 | <0.001 | 0 | T7 | edu | 3.419 | 0.001 | 0.001 |
| T8 | age | -3.982 | <0.001 | 0 | T8 | edu | 3.563 | <0.001 | 0.001 |
| C3 | sex | 1.417 | 0.161 | 0.299 | C3 | amp | -2.238 | 0.027 | 0.055 |
| C4 | sex | 1.348 | 0.178 | 0.299 | C4 | amp | -2.44 | 0.018 | 0.045 |
| Cz | sex | 1.204 | 0.238 | 0.299 | Cz | amp | -1.926 | 0.057 | 0.09 |
| F3 | sex | 1.239 | 0.222 | 0.299 | F3 | amp | -2.501 | 0.016 | 0.045 |
| F4 | sex | 1.305 | 0.197 | 0.299 | F4 | amp | -2.853 | 0.004 | 0.045 |
| F7 | sex | 1.177 | 0.241 | 0.299 | F7 | amp | -2.396 | 0.019 | 0.045 |
| F8 | sex | 1.001 | 0.308 | 0.308 | F8 | amp | -2.227 | 0.029 | 0.055 |
| Fp1 | sex | 1.041 | 0.3 | 0.308 | Fp1 | amp | -2.675 | 0.01 | 0.045 |
| Fp2 | sex | 1.118 | 0.267 | 0.299 | Fp2 | amp | -2.697 | 0.008 | 0.045 |
| Fz | sex | 1.259 | 0.216 | 0.299 | Fz | amp | -2.715 | 0.007 | 0.045 |
| O1 | sex | 1.142 | 0.258 | 0.299 | O1 | amp | -0.941 | 0.345 | 0.345 |
| O2 | sex | 1.374 | 0.173 | 0.299 | O2 | amp | -1.49 | 0.133 | 0.141 |
| P3 | sex | 1.458 | 0.146 | 0.299 | P3 | amp | -2.01 | 0.05 | 0.086 |
| P4 | sex | 1.376 | 0.173 | 0.299 | P4 | amp | -1.778 | 0.079 | 0.106 |
| P7 | sex | 1.359 | 0.178 | 0.299 | P7 | amp | -1.732 | 0.089 | 0.106 |
| P8 | sex | 1.334 | 0.182 | 0.299 | P8 | amp | -1.713 | 0.089 | 0.106 |
| Pz | sex | 1.326 | 0.191 | 0.299 | Pz | amp | -1.844 | 0.068 | 0.1 |
| T7 | sex | 1.2 | 0.234 | 0.299 | T7 | amp | -1.566 | 0.127 | 0.141 |
| T8 | sex | 1.302 | 0.199 | 0.299 | T8 | amp | -2.532 | 0.014 | 0.045 |

**Table S29.** Linear regression between theta SLSW peak-to-peak amplitude and cognitive PD-CRS score. Here, the linear regression controls age, education, and sex, with PD-CRS cognitive score as the dependent variable, and the indicated SLSW feature as the independent variable.

*SLSW delta peak-to-peak amplitude is not associated with cognitive impairment*

| chan | effect | F | p | pfdr | chan | effect | F | p | pfdr |
|---|---|---|---|---|---|---|---|---|---|
| C3 | age | -4.262 | <0.001 | <0.001 | C3 | edu | 3.405 | 0.001 | 0.001 |
| C4 | age | -4.197 | <0.001 | <0.001 | C4 | edu | 3.557 | 0.001 | 0.001 |
| Cz | age | -4.123 | <0.001 | <0.001 | Cz | edu | 3.455 | 0.001 | 0.001 |
| F3 | age | -4.338 | <0.001 | <0.001 | F3 | edu | 3.287 | 0.001 | 0.001 |
| F4 | age | -4.272 | <0.001 | <0.001 | F4 | edu | 3.363 | 0.001 | 0.001 |
| F7 | age | -4.167 | <0.001 | <0.001 | F7 | edu | 3.356 | 0.001 | 0.001 |
| F8 | age | -3.894 | <0.001 | <0.001 | F8 | edu | 3.485 | 0.001 | 0.001 |
| Fp1 | age | -4.103 | <0.001 | <0.001 | Fp1 | edu | 3.368 | 0.001 | 0.001 |
| Fp2 | age | -3.959 | <0.001 | <0.001 | Fp2 | edu | 3.453 | 0.001 | 0.001 |
| Fz | age | -4.237 | <0.001 | <0.001 | Fz | edu | 3.351 | 0.001 | 0.001 |
| O1 | age | -4.003 | <0.001 | <0.001 | O1 | edu | 3.674 | <0.001 | 0.001 |
| O2 | age | -4.095 | <0.001 | <0.001 | O2 | edu | 3.667 | <0.001 | 0.001 |
| P3 | age | -4.063 | <0.001 | <0.001 | P3 | edu | 3.617 | <0.001 | 0.001 |
| P4 | age | -4.199 | <0.001 | <0.001 | P4 | edu | 3.621 | <0.001 | 0.001 |
| P7 | age | -3.911 | <0.001 | <0.001 | P7 | edu | 3.667 | 0.001 | 0.001 |
| P8 | age | -4.057 | <0.001 | <0.001 | P8 | edu | 3.69 | <0.001 | 0.001 |
| Pz | age | -4.19 | <0.001 | <0.001 | Pz | edu | 3.55 | 0.001 | 0.001 |
| T7 | age | -4.019 | <0.001 | <0.001 | T7 | edu | 3.438 | 0.001 | 0.001 |
| T8 | age | -3.897 | <0.001 | <0.001 | T8 | edu | 3.677 | 0.001 | 0.001 |
| C3 | sex | 1.417 | 0.153 | 0.312 | C3 | amp | -2.123 | 0.034 | 0.061 |
| C4 | sex | 1.319 | 0.19 | 0.312 | C4 | amp | -2.155 | 0.032 | 0.061 |
| Cz | sex | 1.18 | 0.242 | 0.312 | Cz | amp | -1.632 | 0.106 | 0.126 |
| F3 | sex | 1.168 | 0.247 | 0.312 | F3 | amp | -2.294 | 0.023 | 0.061 |
| F4 | sex | 1.198 | 0.231 | 0.312 | F4 | amp | -2.457 | 0.015 | 0.059 |
| F7 | sex | 1.074 | 0.295 | 0.323 | F7 | amp | -2.006 | 0.047 | 0.069 |
| F8 | sex | 0.916 | 0.358 | 0.358 | F8 | amp | -2.484 | 0.015 | 0.059 |
| Fp1 | sex | 1.016 | 0.306 | 0.323 | Fp1 | amp | -2.618 | 0.011 | 0.059 |
| Fp2 | sex | 1.072 | 0.29 | 0.323 | Fp2 | amp | -2.69 | 0.008 | 0.059 |
| Fz | sex | 1.201 | 0.234 | 0.312 | Fz | amp | -2.207 | 0.03 | 0.061 |
| O1 | sex | 1.332 | 0.183 | 0.312 | O1 | amp | -1.54 | 0.128 | 0.138 |
| O2 | sex | 1.479 | 0.14 | 0.312 | O2 | amp | -1.758 | 0.08 | 0.101 |
| P3 | sex | 1.493 | 0.138 | 0.312 | P3 | amp | -1.953 | 0.051 | 0.069 |
| P4 | sex | 1.532 | 0.132 | 0.312 | P4 | amp | -2.224 | 0.03 | 0.061 |
| P7 | sex | 1.26 | 0.207 | 0.312 | P7 | amp | -1.504 | 0.131 | 0.138 |
| P8 | sex | 1.404 | 0.163 | 0.312 | P8 | amp | -2.124 | 0.036 | 0.061 |
| Pz | sex | 1.407 | 0.16 | 0.312 | Pz | amp | -2.059 | 0.038 | 0.061 |
| T7 | sex | 1.183 | 0.242 | 0.312 | T7 | amp | -1.412 | 0.16 | 0.16 |
| T8 | sex | 1.22 | 0.234 | 0.312 | T8 | amp | -2.466 | 0.015 | 0.059 |

**Table S30.** Linear regression between delta SLSW peak-to-peak amplitude and cognitive PD-CRS score. Here, the linear regression controls age, education, and sex, with PD-CRS cognitive score as the dependent variable, and the indicated SLSW feature as the independent variable.

*SLSW density is not associated with diurnal somnolence*

| chan | band | F | p | pfdr | chan | band | F | p | pfdr |
|---|---|---|---|---|---|---|---|---|---|
| C3 | Delta | 0.35 | 0.73 | 0.96 | C3 | Theta | -0.75 | 0.45 | 0.79 |
| C4 | Delta | 0.62 | 0.55 | 0.87 | C4 | Theta | -1.24 | 0.22 | 0.79 |
| Cz | Delta | 0.78 | 0.44 | 0.87 | Cz | Theta | -1.53 | 0.13 | 0.79 |
| F3 | Delta | 1.48 | 0.14 | 0.7 | F3 | Theta | -0.01 | 0.99 | 0.99 |
| F4 | Delta | 1.5 | 0.14 | 0.7 | F4 | Theta | -0.77 | 0.44 | 0.79 |
| F7 | Delta | -0.17 | 0.86 | 0.96 | F7 | Theta | -0.18 | 0.85 | 0.99 |
| F8 | Delta | 0.04 | 0.97 | 0.97 | F8 | Theta | -0.78 | 0.43 | 0.79 |
| Fp1 | Delta | 0.22 | 0.82 | 0.96 | Fp1 | Theta | 0.16 | 0.87 | 0.99 |
| Fp2 | Delta | 0.18 | 0.85 | 0.96 | Fp2 | Theta | -0.06 | 0.95 | 0.99 |
| Fz | Delta | 1.21 | 0.23 | 0.85 | Fz | Theta | 0.04 | 0.97 | 0.99 |
| O1 | Delta | 0.79 | 0.43 | 0.87 | O1 | Theta | -0.72 | 0.46 | 0.79 |
| O2 | Delta | 1.11 | 0.27 | 0.85 | O2 | Theta | -0.68 | 0.5 | 0.79 |
| P3 | Delta | 0.73 | 0.47 | 0.87 | P3 | Theta | -1.19 | 0.23 | 0.79 |
| P4 | Delta | 0.91 | 0.36 | 0.87 | P4 | Theta | -1.16 | 0.26 | 0.79 |
| P7 | Delta | 1.45 | 0.15 | 0.7 | P7 | Theta | -0.12 | 0.91 | 0.99 |
| P8 | Delta | 1.63 | 0.11 | 0.7 | P8 | Theta | -0.38 | 0.71 | 0.99 |
| Pz | Delta | 0.54 | 0.59 | 0.87 | Pz | Theta | -1.73 | 0.09 | 0.79 |
| T7 | Delta | 0.58 | 0.56 | 0.87 | T7 | Theta | -0.77 | 0.45 | 0.79 |
| T8 | Delta | 0.1 | 0.92 | 0.97 | T8 | Theta | -0.83 | 0.41 | 0.79 |

**Table S31.** Linear regression between delta and theta SLSW density and diurnal somnolence score (dependent variable). Here, the results of the linear regression, with density as the independent variable, and age, education, and sex as covariates of no interest. No significant result was observed.

*SLSW peak-to-peak amplitude is not associated with diurnal somnolence*

| chan | band | F | p | pfdr | chan | band | F | p | pfdr |
|---|---|---|---|---|---|---|---|---|---|
| C3 | Delta | 0.4 | 0.69 | 0.99 | C3 | Theta | 0.4 | 0.68 | 0.99 |
| C4 | Delta | 0.29 | 0.77 | 0.99 | C4 | Theta | 0.29 | 0.77 | 0.99 |
| Cz | Delta | 0.68 | 0.49 | 0.99 | Cz | Theta | 0.68 | 0.51 | 0.99 |
| F3 | Delta | 0.71 | 0.47 | 0.99 | F3 | Theta | 0.71 | 0.47 | 0.99 |
| F4 | Delta | 0.9 | 0.36 | 0.99 | F4 | Theta | 0.9 | 0.37 | 0.99 |
| F7 | Delta | -0.1 | 0.92 | 0.99 | F7 | Theta | -0.1 | 0.92 | 0.99 |
| F8 | Delta | -0.09 | 0.93 | 0.99 | F8 | Theta | -0.09 | 0.92 | 0.99 |
| Fp1 | Delta | 0.05 | 0.96 | 0.99 | Fp1 | Theta | 0.05 | 0.96 | 0.99 |
| Fp2 | Delta | -0.01 | 0.99 | 0.99 | Fp2 | Theta | -0.01 | 0.99 | 0.99 |
| Fz | Delta | 1.05 | 0.3 | 0.99 | Fz | Theta | 1.05 | 0.3 | 0.99 |
| O1 | Delta | -0.24 | 0.81 | 0.99 | O1 | Theta | -0.24 | 0.82 | 0.99 |
| O2 | Delta | -0.7 | 0.48 | 0.99 | O2 | Theta | -0.7 | 0.48 | 0.99 |
| P3 | Delta | 0.13 | 0.9 | 0.99 | P3 | Theta | 0.13 | 0.89 | 0.99 |
| P4 | Delta | -0.45 | 0.66 | 0.99 | P4 | Theta | -0.45 | 0.66 | 0.99 |
| P7 | Delta | -0.99 | 0.33 | 0.99 | P7 | Theta | -0.99 | 0.32 | 0.99 |
| P8 | Delta | -1.22 | 0.23 | 0.99 | P8 | Theta | -1.22 | 0.23 | 0.99 |
| Pz | Delta | 0.18 | 0.86 | 0.99 | Pz | Theta | 0.18 | 0.86 | 0.99 |
| T7 | Delta | -0.16 | 0.88 | 0.99 | T7 | Theta | -0.16 | 0.87 | 0.99 |
| T8 | Delta | -0.65 | 0.52 | 0.99 | T8 | Theta | -0.65 | 0.52 | 0.99 |

**Table S32.** Results of the linear regression between the diurnal somnolence score (MDS-UPDRS Part 1, item 1.8) as dependent variable, and peak-to-peak amplitude (as independent variable). Age, education, and sex were used as covariates of no interest. No significant result was observed.

*SLSW density is not associated with RBD symptoms*

| chan | term | band | z-value | p | pfdr | band | z-value | p | pfdr |
|---|---|---|---|---|---|---|---|---|---|
| C3 | Group | Delta | -0.677 | 0.499 | 0.928 | Theta | 0.774 | 0.439 | 0.971 |
| C4 | Group | Delta | -0.430 | 0.667 | 0.928 | Theta | 0.418 | 0.676 | 0.971 |
| Cz | Group | Delta | -0.750 | 0.454 | 0.928 | Theta | 1.041 | 0.298 | 0.971 |
| F3 | Group | Delta | -0.657 | 0.511 | 0.928 | Theta | -0.579 | 0.563 | 0.971 |
| F4 | Group | Delta | -0.244 | 0.807 | 0.958 | Theta | 0.064 | 0.949 | 0.971 |
| F7 | Group | Delta | -0.633 | 0.527 | 0.928 | Theta | -0.458 | 0.647 | 0.971 |
| F8 | Group | Delta | 0.089 | 0.929 | 0.978 | Theta | -0.312 | 0.755 | 0.971 |
| Fp1 | Group | Delta | -0.915 | 0.360 | 0.928 | Theta | -1.134 | 0.257 | 0.971 |
| Fp2 | Group | Delta | -0.598 | 0.550 | 0.928 | Theta | -0.659 | 0.510 | 0.971 |
| Fz | Group | Delta | -0.407 | 0.684 | 0.928 | Theta | -0.133 | 0.894 | 0.971 |
| O1 | Group | Delta | -0.447 | 0.655 | 0.928 | Theta | -0.171 | 0.864 | 0.971 |
| O2 | Group | Delta | -0.970 | 0.332 | 0.928 | Theta | 0.854 | 0.393 | 0.971 |
| P3 | Group | Delta | -1.152 | 0.249 | 0.928 | Theta | 0.500 | 0.617 | 0.971 |
| P4 | Group | Delta | -1.310 | 0.190 | 0.928 | Theta | -0.036 | 0.971 | 0.971 |
| P7 | Group | Delta | -0.647 | 0.517 | 0.928 | Theta | 0.397 | 0.691 | 0.971 |
| P8 | Group | Delta | -0.268 | 0.789 | 0.958 | Theta | -0.188 | 0.851 | 0.971 |
| Pz | Group | Delta | -1.269 | 0.204 | 0.928 | Theta | 0.457 | 0.648 | 0.971 |
| T7 | Group | Delta | -0.027 | 0.978 | 0.978 | Theta | -0.423 | 0.673 | 0.971 |
| T8 | Group | Delta | -0.104 | 0.917 | 0.978 | Theta | 0.502 | 0.616 | 0.971 |
| C3 | Density | Delta | -0.362 | 0.717 | 0.921 | Theta | 1.118 | 0.264 | 0.946 |
| C4 | Density | Delta | 0.321 | 0.749 | 0.921 | Theta | 0.268 | 0.789 | 0.946 |
| Cz | Density | Delta | -0.404 | 0.686 | 0.921 | Theta | 1.180 | 0.238 | 0.946 |
| F3 | Density | Delta | -0.647 | 0.517 | 0.921 | Theta | 0.310 | 0.756 | 0.946 |
| F4 | Density | Delta | -0.975 | 0.330 | 0.921 | Theta | 0.651 | 0.515 | 0.946 |
| F7 | Density | Delta | -0.823 | 0.410 | 0.921 | Theta | -0.295 | 0.768 | 0.946 |
| F8 | Density | Delta | -0.221 | 0.825 | 0.922 | Theta | -0.098 | 0.922 | 0.946 |
| Fp1 | Density | Delta | -0.788 | 0.431 | 0.921 | Theta | 0.348 | 0.728 | 0.946 |
| Fp2 | Density | Delta | -0.805 | 0.421 | 0.921 | Theta | 0.405 | 0.686 | 0.946 |
| Fz | Density | Delta | -0.621 | 0.535 | 0.921 | Theta | 0.792 | 0.429 | 0.946 |
| O1 | Density | Delta | -0.072 | 0.943 | 0.988 | Theta | 0.563 | 0.573 | 0.946 |
| O2 | Density | Delta | -0.015 | 0.988 | 0.988 | Theta | -0.879 | 0.379 | 0.946 |
| P3 | Density | Delta | -0.405 | 0.685 | 0.921 | Theta | -0.147 | 0.883 | 0.946 |
| P4 | Density | Delta | -0.563 | 0.574 | 0.921 | Theta | -0.575 | 0.565 | 0.946 |
| P7 | Density | Delta | -0.775 | 0.438 | 0.921 | Theta | 0.136 | 0.891 | 0.946 |
| P8 | Density | Delta | -0.351 | 0.726 | 0.921 | Theta | -0.863 | 0.388 | 0.946 |
| Pz | Density | Delta | -0.285 | 0.775 | 0.921 | Theta | -0.455 | 0.649 | 0.946 |
| T7 | Density | Delta | -1.572 | 0.116 | 0.921 | Theta | 0.068 | 0.946 | 0.946 |
| T8 | Density | Delta | -1.080 | 0.280 | 0.921 | Theta | 0.899 | 0.368 | 0.946 |
| C3 | Interaction | Delta | 1.049 | 0.294 | 0.578 | Theta | -0.470 | 0.638 | 0.999 |
| C4 | Interaction | Delta | 0.787 | 0.431 | 0.608 | Theta | -0.075 | 0.941 | 0.999 |
| Cz | Interaction | Delta | 1.132 | 0.257 | 0.578 | Theta | -0.724 | 0.469 | 0.999 |
| F3 | Interaction | Delta | 1.016 | 0.310 | 0.578 | Theta | 1.005 | 0.315 | 0.999 |
| F4 | Interaction | Delta | 0.585 | 0.558 | 0.663 | Theta | 0.316 | 0.752 | 0.999 |
| F7 | Interaction | Delta | 0.994 | 0.320 | 0.578 | Theta | 0.963 | 0.336 | 0.999 |
| F8 | Interaction | Delta | 0.257 | 0.798 | 0.798 | Theta | 0.792 | 0.428 | 0.999 |
| Fp1 | Interaction | Delta | 1.244 | 0.213 | 0.578 | Theta | 1.559 | 0.119 | 0.999 |
| Fp2 | Interaction | Delta | 0.906 | 0.365 | 0.578 | Theta | 1.062 | 0.288 | 0.999 |
| Fz | Interaction | Delta | 0.759 | 0.448 | 0.608 | Theta | 0.517 | 0.605 | 0.999 |
| O1 | Interaction | Delta | 0.933 | 0.351 | 0.578 | Theta | 0.573 | 0.566 | 0.999 |
| O2 | Interaction | Delta | 1.454 | 0.146 | 0.578 | Theta | -0.496 | 0.620 | 0.999 |
| P3 | Interaction | Delta | 1.606 | 0.108 | 0.578 | Theta | -0.161 | 0.872 | 0.999 |
| P4 | Interaction | Delta | 1.757 | 0.079 | 0.578 | Theta | 0.389 | 0.697 | 0.999 |
| P7 | Interaction | Delta | 1.085 | 0.278 | 0.578 | Theta | -0.001 | 0.999 | 0.999 |
| P8 | Interaction | Delta | 0.698 | 0.485 | 0.615 | Theta | 0.571 | 0.568 | 0.999 |
| Pz | Interaction | Delta | 1.714 | 0.086 | 0.578 | Theta | -0.112 | 0.911 | 0.999 |
| T7 | Interaction | Delta | 0.380 | 0.704 | 0.743 | Theta | 0.847 | 0.397 | 0.999 |
| T8 | Interaction | Delta | 0.437 | 0.662 | 0.740 | Theta | -0.062 | 0.950 | 0.999 |

**Table S33.** Logistic regression between delta and theta SLSW density and the presence/absence of RBD symptoms. Here, the regression controls had age, education, and sex as covariates of no interest, RBD as the dependent variable, and the indicated SLSW feature as the independent variable. An interaction between sub-groups (PD-H, PD-nH) and SLSW density was added to the model. This model was performed only on PD patients. No significant result was observed.

*SLSW peak-to-peak amplitude does not predict RBD symptoms*

| chan | term | band | z-value | p | pfdr | band | z-value | p | pfdr |
|---|---|---|---|---|---|---|---|---|---|
| C3 | Group | Delta | -0.751 | 0.453 | 0.947 | Theta | -0.158 | 0.875 | 0.985 |
| C4 | Group | Delta | -0.488 | 0.626 | 0.947 | Theta | -0.098 | 0.922 | 0.985 |
| Cz | Group | Delta | -0.977 | 0.328 | 0.947 | Theta | -0.248 | 0.804 | 0.985 |
| F3 | Group | Delta | 0.179 | 0.858 | 0.947 | Theta | -0.127 | 0.899 | 0.985 |
| F4 | Group | Delta | -0.067 | 0.947 | 0.947 | Theta | -0.314 | 0.753 | 0.985 |
| F7 | Group | Delta | 0.229 | 0.819 | 0.947 | Theta | 0.197 | 0.844 | 0.985 |
| F8 | Group | Delta | 0.066 | 0.947 | 0.947 | Theta | 0.007 | 0.994 | 0.994 |
| Fp1 | Group | Delta | 0.791 | 0.429 | 0.947 | Theta | 0.298 | 0.765 | 0.985 |
| Fp2 | Group | Delta | 0.401 | 0.688 | 0.947 | Theta | 0.095 | 0.924 | 0.985 |
| Fz | Group | Delta | -0.520 | 0.603 | 0.947 | Theta | -0.441 | 0.659 | 0.985 |
| O1 | Group | Delta | 0.115 | 0.908 | 0.947 | Theta | 0.263 | 0.792 | 0.985 |
| O2 | Group | Delta | -0.129 | 0.897 | 0.947 | Theta | 0.301 | 0.763 | 0.985 |
| P3 | Group | Delta | -0.418 | 0.676 | 0.947 | Theta | -0.096 | 0.923 | 0.985 |
| P4 | Group | Delta | -0.140 | 0.889 | 0.947 | Theta | 0.130 | 0.897 | 0.985 |
| P7 | Group | Delta | -0.220 | 0.826 | 0.947 | Theta | -0.091 | 0.928 | 0.985 |
| P8 | Group | Delta | -0.089 | 0.929 | 0.947 | Theta | 0.131 | 0.896 | 0.985 |
| Pz | Group | Delta | -0.304 | 0.761 | 0.947 | Theta | 0.183 | 0.855 | 0.985 |
| T7 | Group | Delta | 0.165 | 0.869 | 0.947 | Theta | 0.084 | 0.933 | 0.985 |
| T8 | Group | Delta | 0.161 | 0.872 | 0.947 | Theta | 0.408 | 0.683 | 0.985 |
| C3 | Amplitude | Delta | 0.692 | 0.489 | 0.809 | Theta | 0.631 | 0.528 | 0.924 |
| C4 | Amplitude | Delta | 0.785 | 0.433 | 0.809 | Theta | 0.824 | 0.410 | 0.924 |
| Cz | Amplitude | Delta | 0.588 | 0.556 | 0.809 | Theta | 0.545 | 0.586 | 0.924 |
| F3 | Amplitude | Delta | 0.507 | 0.612 | 0.809 | Theta | 0.533 | 0.594 | 0.924 |
| F4 | Amplitude | Delta | 0.567 | 0.571 | 0.809 | Theta | 0.411 | 0.681 | 0.924 |
| F7 | Amplitude | Delta | 0.388 | 0.698 | 0.809 | Theta | 0.413 | 0.680 | 0.924 |
| F8 | Amplitude | Delta | 0.268 | 0.789 | 0.809 | Theta | 0.493 | 0.622 | 0.924 |
| Fp1 | Amplitude | Delta | -0.500 | 0.617 | 0.809 | Theta | 0.040 | 0.968 | 0.968 |
| Fp2 | Amplitude | Delta | -0.275 | 0.783 | 0.809 | Theta | -0.190 | 0.849 | 0.935 |
| Fz | Amplitude | Delta | 0.579 | 0.563 | 0.809 | Theta | 0.305 | 0.761 | 0.935 |
| O1 | Amplitude | Delta | 0.242 | 0.809 | 0.809 | Theta | -0.144 | 0.885 | 0.935 |
| O2 | Amplitude | Delta | 0.940 | 0.347 | 0.809 | Theta | 0.607 | 0.544 | 0.924 |
| P3 | Amplitude | Delta | 0.824 | 0.410 | 0.809 | Theta | 0.917 | 0.359 | 0.924 |
| P4 | Amplitude | Delta | 0.815 | 0.415 | 0.809 | Theta | 0.736 | 0.462 | 0.924 |
| P7 | Amplitude | Delta | 1.005 | 0.315 | 0.809 | Theta | 1.030 | 0.303 | 0.924 |
| P8 | Amplitude | Delta | 1.206 | 0.228 | 0.809 | Theta | 0.686 | 0.493 | 0.924 |
| Pz | Amplitude | Delta | 0.867 | 0.386 | 0.809 | Theta | 0.691 | 0.490 | 0.924 |
| T7 | Amplitude | Delta | 1.104 | 0.270 | 0.809 | Theta | 1.278 | 0.201 | 0.924 |
| T8 | Amplitude | Delta | 0.288 | 0.773 | 0.809 | Theta | -0.185 | 0.853 | 0.935 |
| C3 | Interaction | Delta | 1.178 | 0.239 | 0.866 | Theta | 0.618 | 0.536 | 0.826 |
| C4 | Interaction | Delta | 0.922 | 0.357 | 0.866 | Theta | 0.552 | 0.581 | 0.826 |
| Cz | Interaction | Delta | 1.395 | 0.163 | 0.866 | Theta | 0.681 | 0.496 | 0.826 |
| F3 | Interaction | Delta | 0.229 | 0.819 | 0.866 | Theta | 0.581 | 0.562 | 0.826 |
| F4 | Interaction | Delta | 0.489 | 0.625 | 0.866 | Theta | 0.791 | 0.429 | 0.826 |
| F7 | Interaction | Delta | 0.227 | 0.820 | 0.866 | Theta | 0.290 | 0.771 | 0.826 |
| F8 | Interaction | Delta | 0.429 | 0.668 | 0.866 | Theta | 0.498 | 0.619 | 0.826 |
| Fp1 | Interaction | Delta | -0.276 | 0.782 | 0.866 | Theta | 0.236 | 0.813 | 0.826 |
| Fp2 | Interaction | Delta | 0.123 | 0.902 | 0.902 | Theta | 0.469 | 0.639 | 0.826 |
| Fz | Interaction | Delta | 0.945 | 0.344 | 0.866 | Theta | 0.916 | 0.359 | 0.826 |
| O1 | Interaction | Delta | 0.409 | 0.683 | 0.866 | Theta | 0.337 | 0.736 | 0.826 |
| O2 | Interaction | Delta | 0.641 | 0.521 | 0.866 | Theta | 0.295 | 0.768 | 0.826 |
| P3 | Interaction | Delta | 0.883 | 0.377 | 0.866 | Theta | 0.584 | 0.560 | 0.826 |
| P4 | Interaction | Delta | 0.605 | 0.545 | 0.866 | Theta | 0.367 | 0.713 | 0.826 |
| P7 | Interaction | Delta | 0.768 | 0.442 | 0.866 | Theta | 0.634 | 0.526 | 0.826 |
| P8 | Interaction | Delta | 0.653 | 0.514 | 0.866 | Theta | 0.486 | 0.627 | 0.826 |
| Pz | Interaction | Delta | 0.735 | 0.463 | 0.866 | Theta | 0.288 | 0.774 | 0.826 |
| T7 | Interaction | Delta | 0.360 | 0.719 | 0.866 | Theta | 0.371 | 0.710 | 0.826 |
| T8 | Interaction | Delta | 0.430 | 0.667 | 0.866 | Theta | 0.219 | 0.826 | 0.826 |

**Table S34.** Logistic regression between delta and theta SLSW peak-to-peak amplitude and the presence/absence of RBD symptoms. Here, the regression controls had age, education, and sex as covariates of no interest, RBD as the dependent variable, and the indicated SLSW feature as the independent variable. An interaction between sub-groups (PD-H, PD-nH) and SLSW density was added to the model. This model was performed only on PD patients. No significant result was observed.
